# The pivotal role of the X-chromosome in the genetic architecture of the human brain

**DOI:** 10.1101/2023.08.30.23294848

**Authors:** Zhiwen Jiang, Patrick F. Sullivan, Tengfei Li, Bingxin Zhao, Xifeng Wang, Tianyou Luo, Shuai Huang, Peter Y. Guan, Jie Chen, Yue Yang, Jason L. Stein, Yun Li, Dajiang Liu, Lei Sun, Hongtu Zhu

## Abstract

Genes on the X-chromosome are extensively expressed in the human brain. However, little is known for the X-chromosome’s impact on the brain anatomy, microstructure, and functional network. We examined 1,045 complex brain imaging traits from 38,529 participants in the UK Biobank. We unveiled potential autosome-X-chromosome interactions, while proposing an atlas outlining dosage compensation (DC) for brain imaging traits. Through extensive association studies, we identified 72 genome-wide significant trait-locus pairs (including 29 new associations) that share genetic architectures with brain-related disorders, notably schizophrenia. Furthermore, we discovered unique sex-specific associations and assessed variations in genetic effects between sexes. Our research offers critical insights into the X-chromosome’s role in the human brain, underscoring its contribution to the differences observed in brain structure and functionality between sexes.

**One-sentence Summary:** We investigated the genetic impact of the X-chromosome and the sex differences in the human brain.

---

The genetic foundations governing gene regulation on the X-chromosome are inherently complex due to the XY sex-determination system (*1*). In genetic females, who have two X-chromosomes compared to the single X-chromosome in males, one X-chromosome is inactivated. This inactivation occurs either randomly or with a bias towards a specific parental copy, aiming to balance the transcriptional dosages of X-linked genes between the sexes (*2, 3*). This process, known as X-chromosome inactivation (XCI), facilitates dosage compensation (DC), ensuring that females are cellular mosaics with cells expressing one of the two possible active X-chromosomes (*4*). However, DC is not complete, with only approximately 60-75% of X-linked genes fully silenced (*5*), and this process may vary across different tissues (*6, 7*). As a result, several factors, including DC itself and the differential expression of parent-of-origin genes, significantly influence X-linked gene expression in females (*8*). Notably, in both sexes, the expression levels of genes on the active X-chromosome can be increased to align with those of autosomal genes – a principle known as Ohno’s hypothesis (*9*). This compensatory upregulation of X-linked genes has been observed across various species, including human (*10*) and mouse (*10-12*), indicating a widespread evolutionary strategy for gene expression balance.

The X-chromosome harbors a significant number of genes with predominant expression in brain tissues (*10, 13*), as evidenced by the X-chromosome to autosome expression ratio exceeding one (*10*). This highlights the X-chromosome’s crucial impact on brain anatomy, connectivity, and functionality (*14-17*). Compelling evidence has shown the X-chromosome profoundly influences a wide range of neurological diseases and psychiatric disorders from both genetic and epigenetic viewpoints (*5, 18-20*). For instance, X-linked intellectual disability is a prominently studied brain disorder with 162 related genes identified by 2022 (*21*). Remarkably, the density of genes associated with intellectual disability on the X-chromosome is twice that found on autosomes (*21*). Chromosome mutations, such as sex chromosome aneuploidy, can significantly affect brain structures (*14, 22*), cognitive functions (*23, 24*), behaviors (*24*), and both neurological (*24, 25*) and psychiatric disorders (*26, 27*). A combination of factors, including the high concentration of relevant genes, varying gene expression between sexes, and the influence of epigenetic processes driven by sex steroid hormones, establish the X-chromosome as a nexus for sex differences in the human brain across various age groups (*4, 7, 8, 28-33*). Despite its critical importance, the X-chromosome is frequently overlooked in genome-wide association studies (GWAS), a point underscored by research from Wise et al. (*34*) and Sun. et al. (*35*).

Previous studies have laid foundational work in exploring the genetic contributions of the X-chromosome to brain structures and functions. Smith et al. (*17*) conducted a GWAS analyzing 3,144 complex brain imaging traits in 22,138 subjects. They identified four genome-wide significant loci on the X-chromosome, each with a top single-nucleotide polymorphism (SNP) achieving a p-value less than 7.94 × 10^−12^. They further characterized the genetic loci using expression quantitative trait loci (eQTLs) and examined the genetic co-architectures between brain traits and various health-related conditions. On a different note, Mallard et al. (*16*) examined DC for regional brain measurements, including brain volumes (BVs), cortical thickness (CT), and surface area (SA). Their research highlighted a significant enrichment of X-linked heritability on specific regions of interest (ROIs) within the SA and delved into the genetic basis of these ROIs through an X-chromosome-wide association study (XWAS) (*36*). However, the study by Smith et al. (*17*) was constrained by a smaller discovery sample size than what is now available, while Mallard et al.’s investigation was restricted to brain anatomy. More importantly, there is still a considerable gap in our understanding of the DC profile for brain connectivity and functionality, and the degree to which sex differences in the human brain are modulated by the X-chromosome.

To comprehensively evaluate the X-chromosome’s role in the human brain, we amassed 1,045 complex brain imaging traits derived from structural magnetic resonance imaging (sMRI) for brain anatomical measures, diffusion MRI (dMRI) for white matter (WM) microstructure, and resting-state and task-evoked functional MRI (rfMRI and tfMRI, respectively) for intrinsic and extrinsic brain functions (Table S1). Specifically, we collected 230 sMRI traits encompassing BV, CT, and SA, 635 dMRIs traits covering five diffusion parameters, the fractional anisotropy (FA), mean diffusivity (MD), axial diffusivity (AD), radial diffusivity (RD), and mode of anisotropy (MO) (hereafter DTI traits), and 180 rfMRI and tfMRI traits including the network-level functional amplitude and connectivity. For each diffusion parameter, we derived both the mean value and the top five functional principal components (PCs) within each tract, and the overall average across 21 WM tracts. The tract-mean traits provide average diffusivity metrics of the tracts, while the PC traits capture the finer details and diffusivity heterogeneity within the tract (*37*). Additionally, rfMRI and tfMRI traits were generated using the Glasser360 (*38*) atlas among 12 networks. Further details are provided in the Methods section. The reliability and reproducibility of all traits have been verified in our preceding studies (*37, 39-41*).

An overview of the study design is illustrated in Fig. 1. We first assessed the DC status of each brain imaging trait through model selection, focusing on the narrow-sense heritability associated with the non-pseudo-autosomal region (NPR) SNPs on the X-chromosome via a sex-agnostic analysis. Subsequently, we developed a comprehensive DC atlas for every trait category. In association analysis, we examined both NPR and pseudo-autosomal region (PAR) SNPs on the X-chromosome. Remarkably, we discovered genetic overlaps between the brain imaging traits and various brain-related disorders/phenotypes. Our further explorations delved into sex differences, scrutinizing aspects such as phenotype, heritability, genetic associations, and genetic effects. These explorations have yielded critical insights into the genetic determinants of sex-specific variations in brain structure and functionality. Through this research, we aim to augment the comprehension of the X-chromosome’s contribution to human brain function and development. Our findings will pave the way for enriching future endeavors in biology, clinical sciences, and psychiatry.

**Figure 1:**
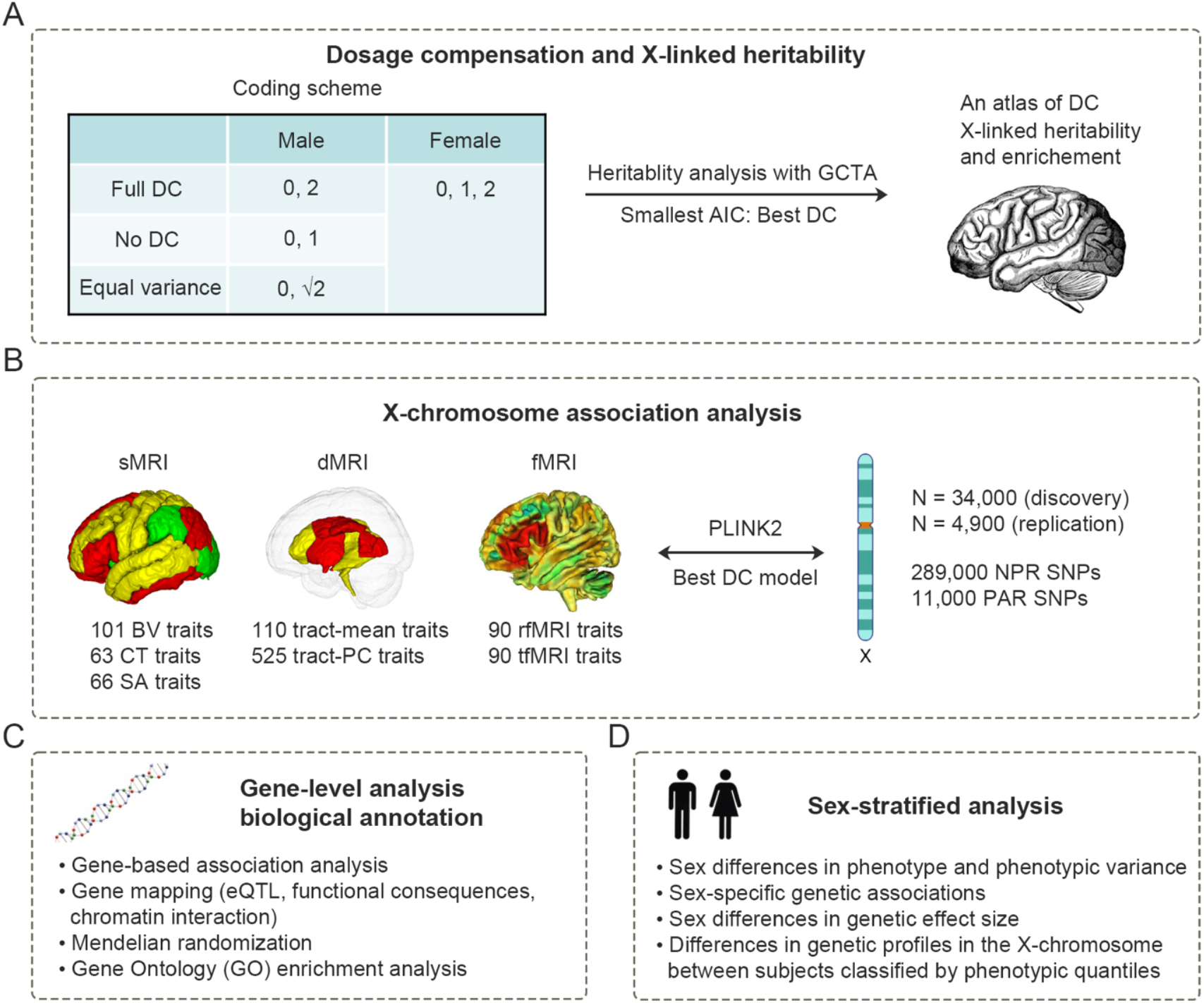
Overview of the study design. This study encompasses four main components. A) We first determined the DC for each trait by evaluating three possible DC models, represented as three different coding schemes. We then established a comprehensive atlas of DC for each trait category. In the meanwhile, we estimated the X-linked heritability and assessed the enrichment. B) Using the optimal DC models, we conducted X-chromosome association analysis across 1,045 complex brain imaging traits for brain anatomy, microstructures, and functionality. C) Through gene-level analysis and biological annotation, we identified associated genes and discovered gene ontology terms with enrichment. We further bridged the association signals with brain-related disorders and other health conditions through gene functional mapping. D) In sex-stratified analysis, we delved into the sex disparities in many aspects, including phenotype, phenotypic variance, X-linked heritability, sex-specific genetic associations, as well as genetic profiles in the X-chromosome between subjects classified by phenotypic quantiles.

## Results

### Dosage compensation in the X-chromosome

We determined a global DC for each trait through model selection. This global DC is not specific to a gene, but reflects an aggregate effect of all the X-linked genes, deriving from the best model to estimate X-linked narrow-sense heritability. Specifically, we utilized three distinct model specifications – full DC, no DC, and equal variance – to jointly estimate the heritability ascribed to autosomes 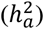 and the heritability credited to the NPR on the X-chromosome 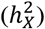 using GCTA (*42*) (Methods). In these models, a female’s genetic profile is always coded as (0, 1, 2), but a male’s genetic profile is coded as (0, 2) for full DC (known as random X-inactivation (*43*)); in no DC, a male’s profile is coded as (0, 1); and in the equal variance model, it is coded as 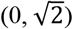. These models illustrate the differing genetic variance between sexes, as males exhibit twice the genetic variance of females in full DC, half in no DC, and identical in the equal variance model. For each trait, the model exhibiting the lowest Akaike information criterion (AIC) was selected as the best model. Given that the heritability linked to the X-chromosome depends on the inactivation status of X-linked loci influencing a trait (*42, 44*), accurately determining the DC status for each trait allows us to more precisely estimate X-linked heritability, as wells as boosting the statistical power of our association analysis.

Our analysis revealed 1,039 out of 1,045 traits (99.4%) exhibited significant total heritability 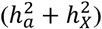, after adjusting for multiple comparisons by controlling the false discovery rate (FDR) at the 0.05 level. Specifically, 819 (78.4%) traits showed a preference for full DC, 187 traits (17.9%) for no DC, and 39 traits (3.7%) for equal variance. Fig. 2A illustrates the DC distribution across each trait category (detailed in Table S2), while Fig. S1 and S2 depict the strength of the preferred DC model for sMRI and DTI tract-mean traits, respectively. The MO, CT (*16*), and AD traits exhibited the highest inclination towards no DC. For traits favoring full DC, no DC, and equal variance, the average 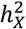 estimates were 1.0% (se = 0.34%), 1.5% (se = 0.63%) and 0.52% (se = 0.44%), respectively (Fig. 2B). The variation in 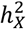 estimates across the three DC categories was statistically significant (Wilcoxon rank sum test, pairwise p-values < 1.8 × 10^−6^). Higher 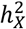 estimates are reasonable for traits favoring no DC, as the lack of XCI leads to an increased gene expression dosage, thereby enhancing the X-chromosome’s influence on genetic regulation.

**Figure 2:**
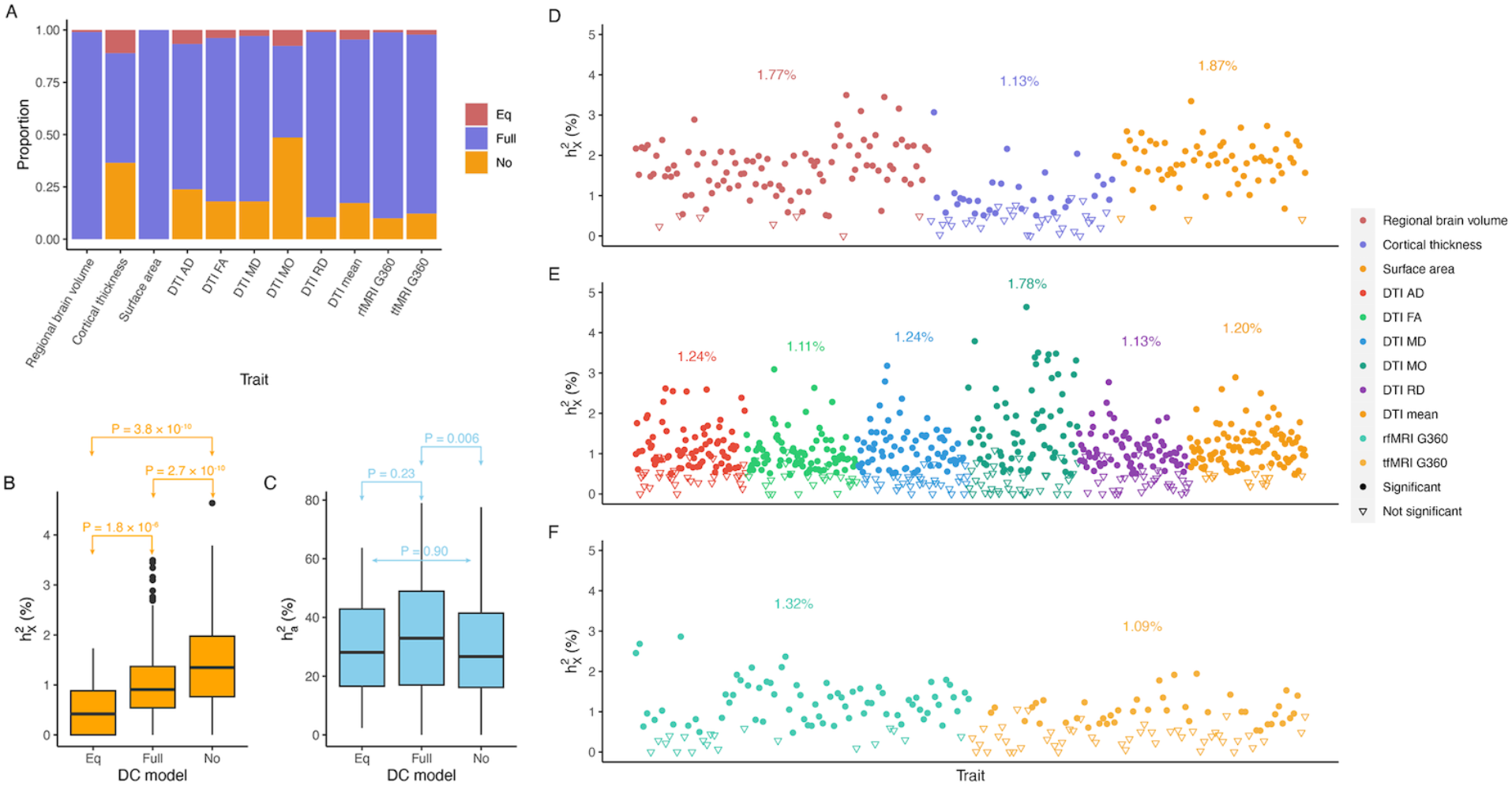
DC and heritability patterns across trait categories. A) Distribution of the three DC groups across trait categories. B)-C) Comparisons of X-linked heritability 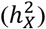 and autosomal heritability 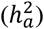 estimates among DC groups, respectively. Unadjusted p-values of the Wilcoxon rank sum test for pairwise comparisons are annotated. D)-E) Scatter plots of 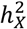 estimates for individual traits in sMRI, DTI, and fMRI, respectively. Different trait categories are color-coded. Traits with significant 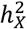 estimates are represented as filled circles, while traits with insignificant 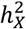 estimates are represented as inverted triangles. The average 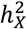 estimate for significant traits is highlighted in its respective color.

Furthermore, our analysis discovered that traits favoring full DC on the X-chromosome exhibited a greater 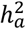 estimate compared to those favoring no DC (Wilcoxon rank sum test, pairwise p-value = 0.006, Fig. 2C). However, no significant difference was observed between traits with full DC and those with equal variance (p-values = 0.22), likely due to the small number of traits with equal variance. Specifically, the average 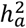 estimate for full DC traits was 34.2%, for no DC traits was 29.7%, and for equal variance traits was 30.1%. This differentiation in 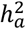 was not attributed to estimation bias, as 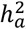 estimates across different DC models were consistent for any given trait. (Fig. S3). The observed differences indicate that 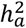 is correlated with the DC status, potentially hinting at autosome-X-chromosome interactions. For example, DNA methylation on autosomes might be *trans* regulated by the X-chromosome (*45*). Moreover, it implies that traits associated with no DC may be more susceptible to non-genetic influences, including environmental factors.

Distinct patterns of DC were discerned across measurements of brain anatomy. Notably, all SA traits favored full DC, while nearly all BV traits showed the same preference, except for the optic chiasm. In contrast, CT traits presented a more varied DC pattern as approximately half of the traits tended towards either no DC or equal variance. While the mean CT still favored full DC, those preferring no DC were distributed across a wide range of cortical regions (Fig. 3A). Despite different preprocessing for raw images, our discoveries resonated with the patterns previously identified by Mallard et al. (*16*). Additionally, existing literature posits that the CT of the motor cortex may be influenced by genes that escape inactivation (*46*). Our findings supported this, as the CT for both the precentral and postcentral cortex (located near the motor cortex) showed patterns consistent with no DC.

**Figure 3:**
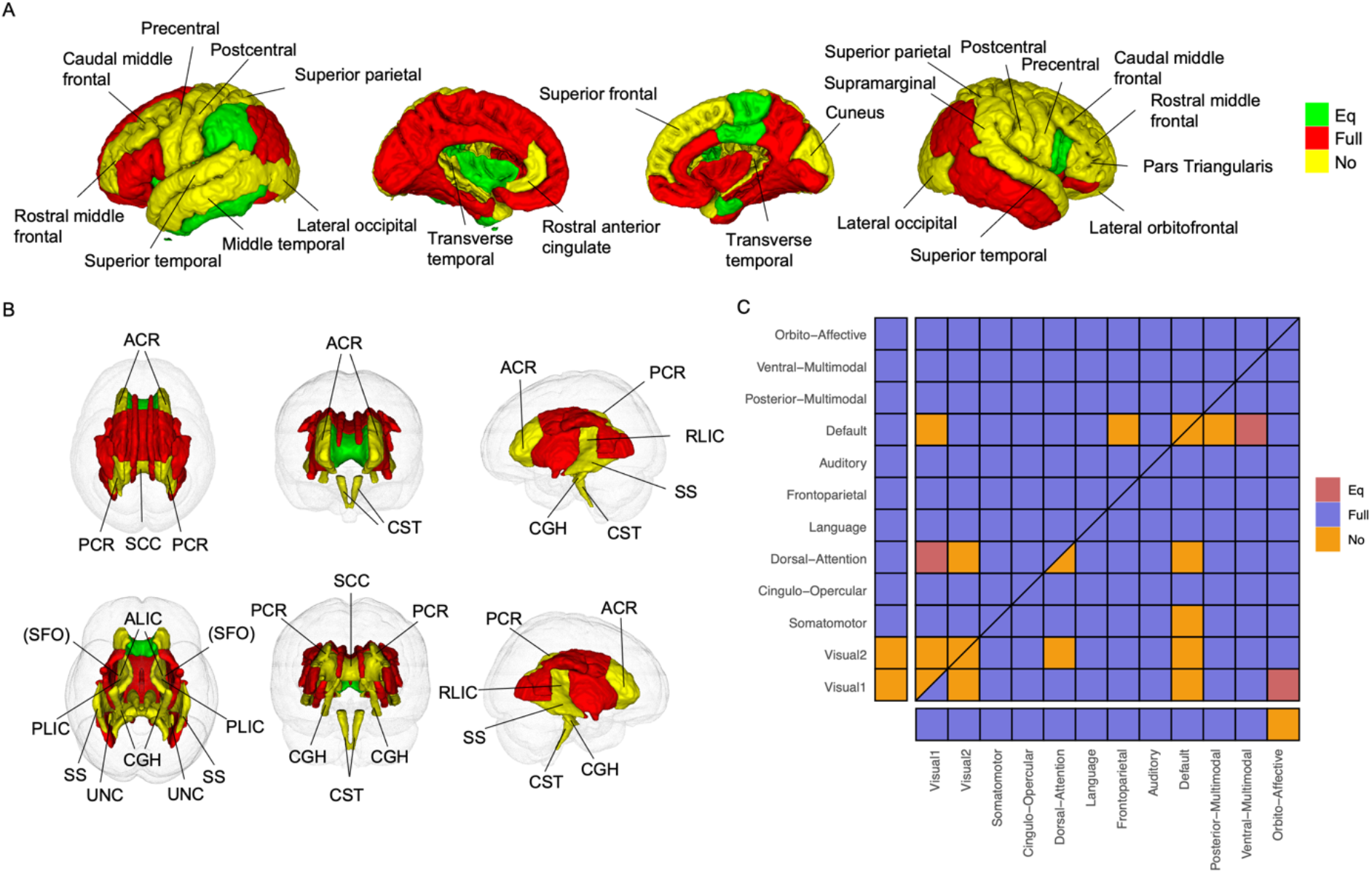
Atlases of DC for CT, MO (tract-mean), and fMRI traits. A) DC atlas for CT: the left two showcase the left hemisphere while the right two display the right hemisphere. Regions of Interest (ROIs) favoring no DC are annotated. B) DC atlas for tract-mean traits assessed by MO shown in six perspectives. The sequence from left to right, top to bottom includes superior, anterior, left, interior, posterior, and right views. Tracts favoring no DC are annotated. “SFO” is in parenthesis since it is blocked by other tracts. C) DC atlas for fMRI traits: The upper triangle illustrates rfMRI, whereas the lower triangle depicts tfMRI. The diagonal, extending from the bottom left to the top right, divides into two sections—the upper triangle portrays DC for rfMRI intra-network connectivity and the lower for tfMRI. Cells on the left margin represent amplitude traits for rfMRI, while those at the bottom indicate amplitude traits for tfMRI.

When assessing WM tracts, MO traits predominantly showed a preference for no DC, whereas RD traits largely favored full DC (Fig. 3B for the DC atlas of MO and Fig. S4 for other DTI metrics). Specifically, MO traits exhibited 51 instances of no DC, while such instances were less than 25 in other metrics (Fig. S5). Notably, all five PCs of MO associated with key WM tracts – the anterior limb of the internal capsule, the corticospinal tract, and the splenium of corpus callosum – uniformly favored no DC (Fig. S5D). This pattern suggests a significant influence of X-escapee genes on microstructures of these tracts.

In our study, rfMRI and tfMRI traits that favored no DC were predominantly associated with the primary and secondary visual networks, as well as the default mode network (Fig. 3C). For instance, the majority of mean connectivity traits, both between and within the visual networks, exhibited a preference for no DC across both rfMRI and tfMRI modalities. We noted that the secondary visual network showed a partial overlap with the superior temporal areas, while the default mode network overlapped with the superior temporal, caudal middle frontal, and rostral middle frontal regions. Intriguingly, CT of these ROIs also favored no DC. Such findings underscore a possible connection between the regional structures of the cerebral cortex and the functional networks.

### X-linked heritability for complex brain imaging traits

We jointly estimated 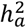 and 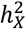 via the GREML analysis of GCTA (*42*) based on the optimal DC model for each trait. Of the total 1,045 traits, 760 (72.7%) traits displayed significant 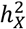 estimates with an average 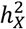 of 1.3% (se = 0.4%) by controlling FDR at the 0.05 level (Fig. 2D-F and Table S2). BV and SA traits predominantly showed high heritability, followed by DTI tract-mean traits and rfMRI traits, whereas tfMRI showcased the fewest proportion (47%) with significant 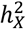 estimates.

We evaluated the enrichment of 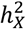 by contrasting the observed 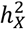 against the expected 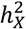 for a genome segment of an equivalent size (Methods). This analysis revealed that 30 traits (2.9%) displayed 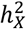 enrichment, while 411 traits (39.3%) exhibited 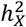 depletion (Fig. S6A-C and Table S2), with the rest showing no significant deviation from expectation. The mean 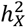 estimate varied significantly across enrichment groups (Wilcoxon rank sum test, pairwise p-values < 4.1 × 10^−8^, Fig. S6D). Notably, SA (hypergeometric test, p-value = 0.0019 < 0.05/22) and rfMRI (p-value = 0.00055) were disproportionately represented in 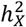 enrichment, while FA (p-value = 7.0 × 10^−7^) and tract-mean traits (p-value = 6.3 × 10^−26^) displayed overrepresentation in depletion. The observed scarcity of traits with enriched 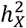 can be attributed to male haploidy and the phenomenon of random XCI in females (*44*), as both factors reduce the genetic variance from the X-chromosome.

The X-chromosome profoundly impacts variations in brain anatomy. Among the 230 traits examined, 179 showed significant 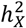 estimates, with an average 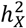 estimate of 1.7% (se = 0.38%). The findings on 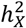 and 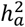 estimates for total BV, mean CT, and total SA echoed results from previous studies (*16, 39, 47, 48*). Interestingly, while no BV traits showed significant enrichment in 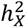, one CT trait (left caudal middle frontal) and seven SA traits were identified as significantly enriched (Fig. S7). The left caudal middle frontal region showcased the highest 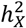 of 3.1% in CT traits, with the X-chromosome accounting for over 14% of total heritability.

Distinctive patterns of 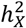 were observed between CT traits and those related to BV and SA. The average 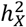 for BV and SA traits were double that of CT traits. Additionally, while the correlations of 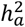 between left and right hemispheres traits were comparable across BV, CT, and SA (Fig. S8A-C), the correlations in 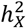 varied significantly. BV traits demonstrated the highest correlation (*r* = 0.81, p-value = 2.1 × 10^−11^), followed by SA traits (*r* = 0.59, p-value = 0.0003) and CT traits showed the lowest correlation (*r* = 0.21, p-value = 0.20) (Fig. S8D-F), despite similar standard errors in 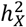 estimates across these anatomical features (*se* = 0.36%∼0.40%). These findings underscore a more pronounced genetic influence of the X-chromosome on BV and SA, with its impact on CT exhibiting greater variability between hemispheres and a more complex DC pattern. This indicates that CT may be subject to unique biological processes during brain development, distinct from those affecting BV and SA (*30, 49*).

The corticospinal tract exhibited distinct DC and heritability enrichment patterns than other WM tracts. It was consistently favoring no DC across all DTI metrics and their PCs. Remarkably, out of the ten functional PC traits enriched in 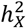, six were associated with the corticospinal tract (Fig. S9). The 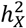 of the fourth PC of the corticospinal tract, evaluated by AD, MD and RD, represented over 80% of the total heritability for these traits, yielding an enrichment ratio nearing 20 (Fig. S6B). This pronounced X-chromosomal influence, coupled with its relatively lower overall heritability compared to other tracts, may contribute to a deeper understanding of the sex differences observed in the development of the corticospinal tract (*50, 51*). Additional findings indicate that an increased dosage from the X-chromosome may significantly reduce the white matter volume within the corticospinal tract(*14*). More extensive analyses regarding the heritability of DTI traits can be found in the Supplementary Text.

While rfMRI and tfMRI exhibited analogous DC patterns, the distribution of 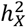 was notably different. The correlation between 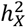 estimates for rfMRI and tfMRI was negaligible (*r* = 0.09, p-value = 0.41), in contrast to a moderate correlation for 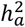 (*r* = 0.34, p-value = 0.001). Specifically, in rfMRI, nine connectivity traits exhibited enrichment in 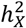, and three of them related to connectivity within the visual networks showcased substantial heritability 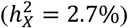, while the corresponding heritability was only 0.5% in tfMRI. Moreover, only three tfMRI connectivity traits related to the language and posterior-multimodal networks were significantly enriched in 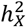 with a mean estimate 1.9%, and these did not overlap with the enriched traits identified in rfMRI (Fig. S10). Taken together, the X-chromosome displayed distinct genetic regulations on the intrinsic and extrinsic brain functions.

### Genetic loci on the X-chromosome associated with complex brain imaging traits

We carried out XWAS on 33,591 subjects from UKB phase 1-3, analyzing 1,045 complex brain imaging traits against 300,000 SNPs on the X-chromosome using PLINK2 (*52*). This included 289,000 NPR SNPs and 11,000 PAR SNPs post-imputation. The specific sample sizes and SNP counts for each trait are detailed in Table S3. For NPR SNPs, we determined the best DC model for each trait following the model selection results. Due to the unavailability of the equal variance model in PLINK2 (where the two male hemizygous genotypes coded as 0 and 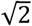), we defaulted to the full DC model (with males coded as 0 and 2) for traits favoring equal variance. Association analysis for PAR SNPs proceeded as with autosomal SNPs. To accurately calibrate the null distribution for correlated test statistics, we employed wild bootstrap technique (*53*), allowing for accurate control of multiple comparisons across the extensive trait analysis (Supplementary Text).

At the genome-wide threshold of 5 × 10^−8^, we identified 29 top NPR SNPs (linkage disequilibrium (LD) *r*^2^ < 0.1), across eight genomic regions (Xp11.4, Xp21.3, Xp22.11, Xp22.12, Xp22.2, Xq13.1, Xq26.3, and Xq28). These SNPs were associated with 72 different brain imaging traits (6 BV, 7 SA, 5 AD, 7 FA, 13 RD, 4 MO, 10 MD, 35 tract-mean, and 1 rfMRI), resulting in a total of 72 trait-locus association pairs (Table S4 and S5, Fig. 4A). The significant associations were disproportionately enriched in the Xq28 region (58 out of 72, proportion test against the proportion of Xq28 in the X-chromosome, p-value < 2.2 × 10^−16^), which was linked to SA and a wide range of tract measures. We did not identify any significant locus in PARs (PAR1 and PAR2 at the two ends of the X-chromosome). Lookups in NHGRI-EBI GWAS catalog (*54*) revealed 29 novel trait-locus association pairs (Table 1). We found evidence that some top SNPs were related to brain structures in previous studies. For instance, rs2272737 was associated with a variety of DTI measures (*17*); rs12843772 was associated with BV, cortical areas (*17*), and brain shape (*55*).

**Table 1:** Novel trait-locus pairs on the X-chromosome for complex brain imaging traits in sex-agnostic analysis. The identified trait-locus pairs are compared to results on the NHGRI-EBI GWAS catalog data (up to June 2023). A pair is considered novel if either the trait is novel at the locus, or the locus is novel for the trait. The full names of white matter tracts are present in Table S1. The raw p-values are adjusted by using the wild bootstrap approach. The physical locations of loci are determined based on GRCh37.

| Trait | UniqID | RsID | p-Value | p-Value (adj) | Start | End | Region |
| --- | --- | --- | --- | --- | --- | --- | --- |
| Total SA (left) | 23:154740180:A:G | rs76536573 | 3.09E-13 | 1.86E-12 | 153732991 | 154949953 | q28 |
| Total SA (right) | 23:154678628:C:G | rs28880039 | 1.13E-11 | 2.73E-10 | 153732991 | 154949953 | q28 |
| SA of lateral occipital (left) | 23:154440161:C:T | rs11156600 | 2.14E-12 | 2.74E-11 | 154018581 | 154949953 | q28 |
| SA of superior frontal (left) | 23:136567864:C:T | rs5931154 | 1.50E-10 | 9.12E-09 | 136500146 | 137073809 | q26.3 |
| SA of supramarginal (left) | 23:136505579:C:T | rs2743914 | 8.02E-15 | 1.10E-14 | 136471150 | 137073809 | q26.3 |
| SA of supramarginal (right) | 23:136500146:A:G | rs2840674 | 4.65E-12 | 8.01E-11 | 136450688 | 136841801 | q26.3 |
| BV of insula (left) | 23:11381616:A:G | rs5935037 | 2.83E-10 | 1.94E-08 | 11381616 | 11594043 | p22.2 |
| BV of insula (right) | 23:11381616:A:G | rs5935037 | 1.28E-10 | 6.65E-09 | 11381616 | 11594043 | p22.2 |
| BV of cerebrospinal fluid | 23:38009121:A:G | rs35318931 | 6.71E-12 | 9.84E-11 | 37933437 | 38017972 | p11.4 |
| BV of thalamus proper (left) | 23:28140326:A:G | rs2131673 | 4.69E-14 | 1.32E-13 | 28033828 | 28284470 | p21.3 |
| BV of thalamus proper (right) | 23:28140326:A:G | rs2131673 | 1.99E-13 | 1.01E-12 | 28033828 | 28284470 | p21.3 |
| BV of ventral DC (right) | 23:68377485:A:G | rs2361468 | 4.22E-11 | 8.43E-09 | 68354618 | 68384580 | q13.1 |
| PCR (AD PC1) | 23:152589891:A:G | rs5970440 | 4.17E-10 | 3.60E-08 | 152576561 | 152656246 | q28 |
| PLIC (AD PC1) | 23:152590165:C:G | rs5970442 | 3.20E-10 | 2.53E-08 | 152576561 | 152656246 | q28 |
| PTR (AD PC3) | 23:154740180:A:G | rs76536573 | 5.57E-13 | 6.00E-12 | 154379088 | 154972517 | q28 |
| SCR (AD PC1) | 23:152590165:C:G | rs5970442 | 1.41E-12 | 1.53E-11 | 152576561 | 152656246 | q28 |
| BCC (FA PC3) | 23:39418196:C:T | rs57718759 | 1.23E-10 | 7.02E-09 | 39383773 | 39494923 | p11.4 |
| UNC (FA PC1) | 23:152908152:A:G | rs2301142 | 1.77E-13 | 8.88E-12 | 152877997 | 152908887 | q28 |
| BCC (RD PC2) | 23:39464147:C:T | rs7888246 | 4.66E-12 | 8.02E-11 | 39383773 | 39494923 | p11.4 |
| PCR (MO PC1) | 23:152589891:A:G | rs5970440 | 9.85E-10 | 9.08E-09 | 152576561 | 152656246 | q28 |
| PLIC (MO PC4) | 23:150542930:G:T | rs201850695 | 1.10E-13 | 4.22E-12 | 150497978 | 150614462 | q28 |
| PLIC (MO PC5) | 23:150514840:C:T | rs5924909 | 2.75E-10 | 1.25E-09 | 150514840 | 150614316 | q28 |
| SCR (MO PC1) | 23:152638744:G:T | rs67596711 | 1.25E-11 | 3.12E-10 | 152576561 | 152656246 | q28 |
| BCC (FA mean) | 23:152906292:A:T | rs3761536 | 1.16E-10 | 2.91E-08 | 152886255 | 152908887 | q28 |
| PTR (FA mean) | 23:152898261:A:G | rs4328011 | 3.27E-10 | 2.60E-08 | 152894551 | 152907479 | q28 |
| UNC (FA mean) | 23:152908152:A:G | rs2301142 | 1.22E-12 | 1.01E-10 | 152877997 | 152908887 | q28 |
| SCR (AD mean) | 23:152590019:C:T | rs5970441 | 1.10E-10 | 6.02E-09 | 152576561 | 152648336 | q28 |
| SCR (MO mean) | 23:152638744:G:T | rs67596711 | 2.12E-11 | 6.41E-10 | 152576561 | 152656246 | q28 |
| Amplitude of the language network (rfMRI) | 23:19937727:A:C | rs7880364 | 1.54E-10 | 4.12E-08 | 19572473 | 20322238 | p22.12 |

**Figure 4:**
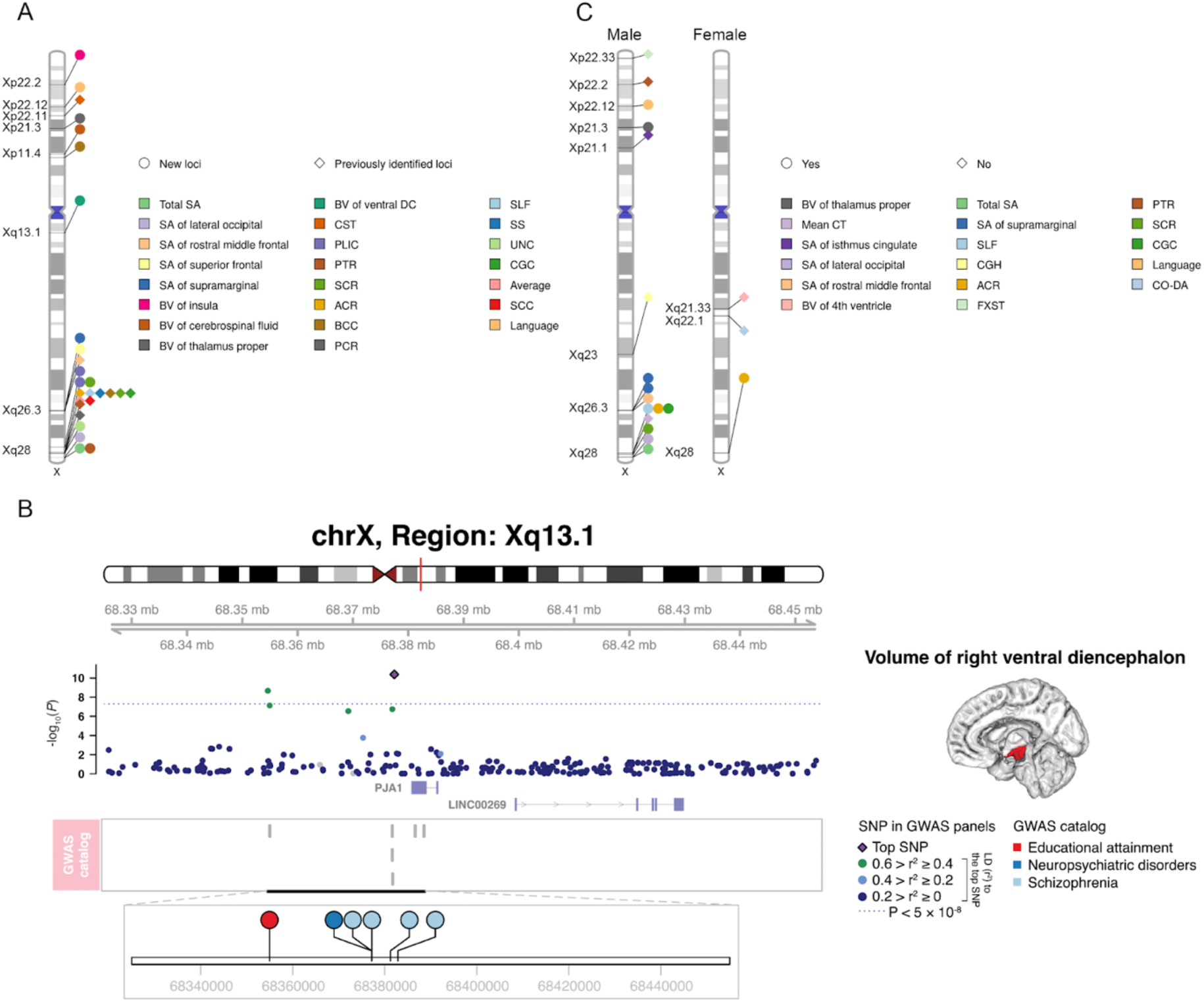
Main results in X-chromosome association analysis. A) Genomic loci associated with complex brain imaging traits identified in sex-agnostic association analysis. Ideogram illustrating the genomic regions affecting brain imaging traits. Each trait is represented by a unique color, with the corresponding genomic region annotated directly on the ideogram. The signal point for white matter tracts indicates that at least one of the 25 related traits (20 PC traits and five mean traits) is associated with the genomic region. Novel identifications are represented in circles, while replicating for previous discoveries are represented in diamonds. B) A selected genetic locus with important colocalizations with brain-related phenotypes. The top SNP, defined as the SNP with the lowest p-value in the locus, surpasses the 5 × 10^−8^ threshold after adjusting for multiple comparisons using the wild bootstrap approach. However, the p-values shown in the figure remain unadjusted. The top SNP along with all the SNPs in LD (*r*^2^ > 0.6) in the same locus are the targets for XWAS results lookup in the NHGRI-EBI GWAS catalog (up to June 2023). The volume of the right ventral diencephalon exhibits shared genetic links with educational attainment and schizophrenia, pinpointed at an Xq13.1 locus by rs2361468. C) Genomic loci associated with complex brain imaging traits identified in sex-stratified association analysis. The label “Language” denotes the mean amplitude trait of the language network in rfMRI. “CO-DA” stands for the mean connectivity trait between the cingulo-opercular and dorsal-attention networks in rfMRI. Trait-locus pairs that are overlapped with those identified in the sex-agnostic analysis are represented as circles, while those specific to sex-stratified analysis are represented as diamonds.

To ensure the robustness of our XWAS findings, we verified the XWAS results from two aspects considering the unique features of the X-chromosome. First, we estimated genetic effects separately for males and females in a sex-stratified analysis, followed by a meta-analysis via METAL (Methods). The majority (63 out of 72) of significant trait-locus pairs were also discovered in the meta-analysis, with three additional ones emerging exclusively through the meta-analysis (Table S6). Second, we conducted replication XWAS analysis using separate datasets in UKB: phase 4 subjects with European ancestry (UKBE, n = 4,181), phase 1-4 subjects with South Asian ancestry and Chinese ancestry (UKBSAC, n = 462), and phase 1-4 subjects with African ancestry (UKBA, n = 295), to validate our discoveries (Table S7). All 72 index SNPs displayed consistent effect sizes between discovery and replication studies using UKBE (proportion test against 0.5, p-value < 2.2 × 10^−16^). Out of 72 index SNPs, 10 (13.9%) were replicated at a conservative Bonferroni threshold (p-value < 0.05/72 = 0.0007). However, replication was not achieved for the UKBSAC and UKBA groups, likely due to their smaller sample sizes.

Further meta-analysis across these ethnic groups (total n = 4,938) allowed for the replication of 23 (31.9%) index SNPs at the same Bonferroni threshold, with the effect direction for each replicated SNP remaining consistent across ancestry groups. Finally, we meta-analyzed the XWAS results of phase 1-3 and phase 4 for European subjects (n = 37,772) and identified 104 significant trait-locus pairs. Among them, 33 novel associations were not identified in the discovery phase, highlighting new genomic regions Xp11.23 and Xq21.1 (Table S8).

### Shared genetic architectures with brain-related disorders and other phenotypes

The identified loci constructed genetic connections between complex brain imaging traits and a broad spectrum of neurological diseases, psychiatric disorders, and cognitive functions, among others (Table S9). Notably, the volume of the right ventral diencephalon (known as hypothalamus) was significantly associated with rs2361468 at Xq13.1, where it tagged numerous variants in LD that have been linked to schizophrenia (*56-59*). Additionally, another SNP rs62606709 at the same locus has been associated with educational attainment (*60*) (Fig. 4B). The Xq13.1 region is enriched with variants implicated in neuroticism (*18*) and a variety of neuropsychiatric disorders, including schizophrenia (*61, 62*), autism spectrum disorder (ASD) (*62, 63*), bipolar disorder (BD) (*62*), major depressive disorder (MDD) (*62*) and Parkinson’s disease (PD) (*64*). Through further literature review, we found more evidence supporting the association between the volume of the ventral diencephalon and various neuropsychiatric and neurological disorders. For instance, ventral diencephalon is implicated in many pathways found disrupted in schizophrenia (*65*) and enlargement of ventral diencephalon was observed in patients with schizophrenia (*66*) and MDD (*67*). Conversely, atrophy in the ventral diencephalon has been linked to AD (*68, 69*), MDD (*70-72*), late-life depression (*73*), PD (*74*), and spinocerebellar ataxias (SCA3) (*75*).

Apart from brain-related disorders, we also identified colocalizations between brain imaging traits and various conditions and health-related traits (Table S9). In Xq26.3, the genetic loci associated with SA of supramarginal influenced the onset of myopia (*76*) and refractive error (*77*). Meanwhile, the genetic loci associated with DTI traits in Xq28 were connected to testosterone levels (*78, 79*), type 2 diabetes (*80-84*), blood-related traits including hemoglobin (*85*), hematocrit (*85*), red blood cell count (*85*), serum uric acid levels (*86*), serum creatinine levels (*86*), blood urea nitrogen levels (*86*), and factor VIII levels (*87*), as well as cardiovascular disorders, such as venous thromboembolism (*88*). These discoveries highlighted the X-chromosome’s significant role in influencing both brain-related and systemic health outcomes.

### eQTL mapping and biological annotation

We conducted eQTL mapping to investigate the functionality of the significant independent SNPs (LD *r*^2^ < 0.6) and focused on the connections with neuropsychiatric disorders (Methods, Table S10). External eQTL datasets from the Genotype-Tissue Expression (*89*) (GTEx v8) and CommonMind Consortium (*20*) (CMC) were employed. This analysis underscored significant eQTLs in Xq28 associated with schizophrenia. Notably, rs4370701, an eQTL for *FAM3A* affecting gene expression in the cerebellum, cortex, and hypothalamus, was associated with the left total SA. Mutations in *FAM3A* have been linked to schizophrenia (*58*). Additionally, our analysis reaffirmed the association between rs2361468 at Xq13.1 and the volume of the right ventral diencephalon. This variant was also found to regulate gene expression of *PJA1*, whose mutations have been implicated in increasing the risk for schizophrenia (*59*) and other neuropsychiatric disorders (*62*). To further explore the genetic associations from different aspects, we conducted additional gene-based and gene-set analysis, and gene mapping based on functional consequences using FUMA (Methods). The detailed results are presented in the Supplementary Text.

Through eQTL mapping, we have linked the significant variants to gene expression levels, but the causal influence of gene expression on the brain traits remained ambiguous. We employed summary statistics-based Mendelian randomization (SMR) (*90*) to investigate whether XWAS traits could be modulated by gene expression. We further used the HEIDI test (*90*) to distinguish pleiotropy of causal variants from linkage (Methods). The eQTL data containing NPR SNPs across 1,639 probes was incorporated from Sidorenko et al.’s (*44*) CAGE whole-blood analysis. Upon adjusting for FDR at the 0.05 level, 14 genes were identified as having potential causal effects on the structural and functional brain traits (Table S11). Applying the threshold p-HEIDI > 0.05 (*90*) to filter out linkage from pleiotropy of causal variants, 12 associations indicative of SNPs that concurrently influence gene expression and brain traits.

Among these, five genes identified through eQTL mapping – *FAM3A, PJA1, TMLHE, PLXNA3*, and *ZNF275* – were also found to have causal impacts on the brain imaging traits. We have previously illustarted the connections between *FAM3A, PJA1* and schizophrenia. Additionally, the nondysmorphic autism-linked gene *TMLHE* (*91, 92*) exerted a causal effect on AD of the posterior thalamic radiation, the BV of the right ventral diencephalon, and multiple SA traits. The ASD-linked gene *PLXNA3* was found to influence the MO of the posterior corona radiata (*93*). These findings reinforce the genetic interplay between schizophrenia, autism, and brain imaging traits.

Finally, we conducted biological annotation using the DAVID Bioinformatics Database (*94*) and SynGO on all the prioritized genes identified by FUMA (Methods). Within DAVID, 182 genes underwent cataloging and subsequent analysis (Table S12). At the FDR threshold of 0.05, the genes were enriched in transcription elongation (IPR021156 and PIRSF008633) and were associated with diseases such as autism (KW-1269) and intellectual disability (KW-0991). At the nominal significance level (p-value < 0.05), these genes were enriched in biological pathways, including “axon development” (GO:0061564), “neurogenesis” (UP_KW-0964), and “nervous system development” (GO:0007399). Through SynGO analysis (Fig. S11 and Table S13), 14 genes were uniquely mapped to SynGO-annotated genes, and 11 genes received cellular component annotations: four were postsynaptic, two as both pre- and postsynaptic, three as presynaptic, and two could not be mapped to any specific compartment.

### Disparity of genetic associations between sexes

We uncovered substantial differences between sexes across a wide array of brain imaging traits (Table S14). A striking 88.0% of analyzed traits showed significant phenotypic sex differences after adjusting for the FDR level of 0.05. More detailed results for distribution of phenotypic differences across imaging modalities and the relationship between DC, 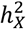 and phenotypic differences are presented in the Supplementary Text. Further, in the sex-stratified analysis of heritability (Methods), 155 traits demonstrated significant 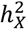 estimates for males compared to only two traits for females (Fig. S12-13 and Table S15). These findings suggest that the observed disparities in brain traits between males and females could be partially attributable to genetic regions on the X-chromosome that are uniquely associated with one sex.

To verify this hypothesis, we conducted sex-stratified XWAS for 16,094 males and 17,558 females on the 1,045 brain imaging traits. Due to reduced sample sizes post-split, we used a significance threshold of 1.0 × 10^−8^ for all analyses in this section, aligning with Bernabeu’s (*95*) approach in a related study. In the NPR for males, we identified 25 trait-locus pairs across seven genomic regions, impacting 25 distinct traits (Fig. 4C and Table S16). Interestingly, 20 out of these traits displayed significant sex differences. However, the female-specific NPR analysis yielded only five trait-locus pairs across three genomic regions, affecting five traits (Fig. 4C), with each manifesting pronounced sex disparities. In the PAR for males, we found a trait-locus pair in Xp22.33 related to FA of the fornix and stria terminalis. However, no findings emerged from the female-specific analysis.

Substantial discrepancies were evident in association patterns between males and females. Xq28 emerged as the only genomic region where both male and female-specific associations were identified (Fig. 4C). The only shared traits between sexes in Xq28 were related to the anterior corona radiata. A notable 80% (20 of the 25) of the male NPR trait-locus pairs were also identified in the sex-agnostic XWAS, compared to 60% (three out of five) for females (Fig. 4C). This suggests that the significant loci detected in the sex-agnostic XWAS are primarily driven by male-associated genetic variations. Most traits with significant loci (regardless of sex) favored full DC. The paucity of significant loci in females aligns with the observation that, for full DC-favoring traits, more genetic variance manifests in males than in females, which highlights the necessity for XWAS statistical methodologies to adequately account for heteroskedasticity between sexes.

To maximize the insights from our data, we performed a meta-analysis of the sex-stratified XWAS, incorporating UKB phase 1-3 and phase 4 subjects of European ancestry, resulting in final sample sizes of 18,025 males and 20,054 females (Table S17). This analysis saw the number of male-specific trait-locus pairs in NPR increase from 31 to 61 at our chosen significance threshold, while the female dataset gained an additional six pairs in NPR. In PAR, the male-specific association became insignificant, and no more pairs were observed for females.

Genetic variants can have starkly different effects between sexes, and the extent of these differences can vary across traits and tissues (*95, 96*). The variations might illuminate the observed disparities in human brain structures across sexes (*4, 8, 32, 97*). We pinpointed three trait-locus pairs across three genomic regions that exemplify these sex differences (Fig. S14 and Table S18). All the traits displayed significant sex differences, including CT of left rostral middle frontal, as well as FA and RD of the anterior corona radiata. All the variants had moderate effect sizes but distinct directions between sexes, and therefore none of the trait-locus pairs overlapped with sex-specific associations. For example, rs12387759 in Xq27.3 revealed differences between sexes on CT of the left rostral middle frontal. Similarly, rs62589244 in Xp11.4 had divergent effects on RD of the anterior corona radiata.

Differences in genetic effects imply an interaction between genetic variants and sex. Traditional XWAS which primarily focus on the additive effect, may overlook these variants with varying effect directions in a linear model, leading to the “masking of genetic effect” problem (*95*). Through meta-analysis using Stouffer’s method (*98*), we identified variants overlooked in the sex-agnostic XWAS (Fig. S14). All the above trait-locus pairs with varying genetic effects were significant. This highlights the need for XWAS to account for interactions between variants and sex, reflecting XCI uncertainty (*43*).

Finally, we hypothesized that there might be distinct genetic profiles in the X-chromosome for subjects that were consistently located at two tails of phenotypic distributions across imaging modalities. Segregating these subjects by sex, we compared their genetic profiles of the X-chromosome using Fisher’s exact test (Methods). We identified 29 significant genetic loci for males (p-value < 1.0 × 10^−8^), spanning the entire X-chromosome without clustering in specific regions. Some of these loci were consistent with findings from our sex-stratified analysis, such as a locus related to CT at Xq28 (Fig. S15 and Table S19). In contrast, no significant results were observed for females. This surprising outcome suggests that brain phenotypic variations in males can be reflected by diverse X-chromosome genetic profiles, whereas other factors might drive brain variations in females.

## Discussion

Our study delved into the significant role of the X-chromosome in influencing brain anatomy, microstructure, and functionality by examining 1,045 complex brain imaging traits in UKB with sample size 38,529. We assessed three DC models: full DC, no DC, and equal variance in GCTA-GREML (*42*), to determine the most appropriate DC status for each trait. This analysis aimed to reflect the DC behaviors of effective genes during development. Our findings contribute to a detailed atlas of DC and underscore the enrichment of X-linked heritability across brain imaging traits. We identified 29 novel trait-locus pairs in the NPR using the wild bootstrap approach for more accurately correcting multiple hypothesis testing. This research highlights the genetic co-architectures between brain measurements and various brain-related disorders, particularly schizophrenia. Furthermore, we discovered sex-specific genetic association patterns and differing genetic effects between sexes, and the NPR SNPs on the X-chromosome accounted for more phenotypic variations in males.

We discovered intriguing interactions between the X-chromosome and autosomes in the human brain. Specifically, we observed that traits favoring no DC displayed significantly higher X-linked heritability but reduced autosomal heritability compared to traits with full DC. For no DC traits, the X-chromosome contributed to 7.1% of the total heritability, contrasting with the 3.8% contribution for full DC traits. We speculate that no DC traits may be significantly influenced by complex transcriptional, regulatory, and epigenetic processes during brain development (*31, 33*). For example, sex steroid receptors often signal through epigenetic actions (*99*). Several epigenetic mechanisms, such as the levels of DNA methylation and acetylation, are sex-specific in the brain (*100*). Recent studies have demonstrated that sex chromosomes can also induce sex differences in somatic gene expression in the absence of hormonal differences (*29*). Further evidence indicates that some genes escaping XCI produce proteins that regulate chromatin structures, potentially influencing autosomal gene expression levels (*29*). This includes the histone demethylases *UTX* and *KDM5C* (*101, 102*), the histone deacetylase 8 (*103-105*), and the histone acetyltransferase complex subunits, such as male-specific lethal 3 (*106*) and mortality factor 4-like 2 (*107*). Such *trans*-modifications, which do not alter the nucleotide sequence, might not be reflected in the narrow-sense heritability, accounting for the reduced heritability ascribed to autosomes. In essence, our analysis offers insights into potential XCI escape at the trait level in the human brain.

The Xq28 genomic region was overly represented in association signals as it contains more than 40% of the identified protein-coding genes. A third of WM tracts were linked to a compact 40 Kb segment within Xq28 (152,876,000∼152,916,000, GRCh37). The importance of Xq28 is further underscored by its connection to various intellectual and developmental disabilities. For example, mutations in the *CLIC2* and *VBP1* genes, located in the int22h-1/int22h-2-mediated duplication area of Xq28, have been implicated as potential contributors to these disorders (*108-110*). Additionally, loss-of-function mutations in the *MECP2* gene are associated with Rett syndrome (*111*), which mainly affects brain development in girls. In males, these mutations lead to a range of clinical outcomes, from mild intellectual challenges to severe neonatal encephalopathy, and in some cases, premature death (*112*). Considering the substantial role of Xq28 in influencing brain structure and function, coupled with its connection to a wide spectrum of brain disorders, future research should focus on characterizing the transcriptomic, proteomic, and epigenetic profiles for this region to fully understand the underlying biological processes.

In addition to sex-agnostic analyses, we systematically explored sex disparities in human brain measurements, including phenotype, phenotypic variance, X-linked heritability, and genetic associations. We observed males exhibited greater phenotypic variance and X-linked heritability across most traits, as well as a higher number of sex-specific genetic associations and better alignment between genetic profile and phenotypic variation. These findings reflect the biological process where one of the X-chromosomes in females is randomly silenced in most tissues and cells, potentially contributing to the observed differences. Furthermore, we discovered that genetic effects of certain variants significantly diverged between sexes, indicating interactions between sex and variants. Combined with many colocalizations between brain measurements and sex-hormone related traits, such as testosterone levels and sex hormone binding globulin (SHBG), we speculate that sex hormones mediated the genetic effect on human brain development and sexual dimorphism. However, verification of the hypothesis requires more data of transcriptome, proteome, and metabolome (termed as “multi-omics” (*113*)), which is currently barren for the X-chromosome. We earmark this line of inquiry for future research, emphasizing the necessity for both innovative methodologies and robust multi-omics data dedicated to the X-chromosome.

## Data Availability

The individual-level data used in the present study have been openly available from the UK Biobank (https://www.ukbiobank.ac.uk/) study.

## Acknowledgements

We thank the participants in the UKB study for their contribution and the research teams for their work in collecting, processing, and disseminating these datasets for analysis. We thank University of North Carolina at Chapel Hill, and the Research Computing groups for providing computational resources and support that have contributed to the research results.

## Author contributions

Conceptualization: ZJ, BZ, HZ, LS

Methodology: ZJ

Preprocessing: TFL, XW, TYL, YY, HS, PYG, JC

Investigation: ZJ

Visualization: ZJ

Funding acquisition: HZ, BZ, TFL, YL

Project administration: HZ

Supervision: BZ, LS, HZ

Writing – original draft: ZJ

Writing – review & editing: ZJ, PFS, YL, JLS, DL, LS, HZ, TFL

## Data and materials availability

This research was conducted using the UKB resource (application number 22783), subject to a data transfer agreement. The UKB has obtained ethics approval from the Northwest Multi-Centre Research Ethics Committee (MREC, approval number: 11/NW/0382), and obtained written informed consent from all participants prior to the study. The eQTL summary statistics from the CAGE whole-blood study can be downloaded at https://cnsgenomics.com/content/data. All files to generate annotation used in H-MAGMA can be accessed at https://doi.org/10.5281/zenodo.5503876. The summary statistics (*114*) generated by association analyses in the current study can be accessed at https://bigkp.org/ and Zenodo (https://zenodo.org/record/8317844). All software and packages used in this study are publicly available: PLINK2 (www.cog-genomics.org/plink/2.0/), METAL (https://genome.sph.umich.edu/wiki/METAL), GCTA (https://yanglab.westlake.edu.cn/software/gcta/#Overview), DAVID Bioinformatics DataBase (https://david.ncifcrf.gov/home.jsp), SynGO (https://syngoportal.org/), FUMA (https://fuma.ctglab.nl).

## Supplementary Materials

Materials and Methods

Supplementary Text

Figs. S1 to S21

Tables S1 to S23

References (115-142)

