## supplementary_materials for "The pivotal role of the X-chromosome in the genetic architecture of the human brain"

Science
AAAS

Supplementary Materials for

### The pivotal role of the X-chromosome in the genetic architecture of the human brain

**Authors:** Zhiwen Jiang, Patrick F. Sullivan, Tengfei Li, Bingxin Zhao, Xifeng Wang, Tianyou Luo, Shuai Huang, Peter Y. Guan, Jie Chen, Yue Yang, Jason L. Stein, Yun Li, Dajiang Liu, Lei Sun, and Hongtu Zhu*

**The PDF file includes:**

Materials and Methods
Supplementary Text
Figs. S1 to S21
Captions of Tables S1 to S23
References 115-142

**Other Supplementary Materials for this manuscript include the following:**

Tables S1 to S23

#### Materials and Methods

Image acquisition and processing

The structural MRI (sMRI), diffusion MRI (dMRI), resting-state functional MRI (rfMRI), and task-evoked functional MRI (tfMRI) images were obtained from the UK Biobank (http://www.ukbiobank.ac.uk/resources/) with application 22783. These images have been subject to appropriate preprocessing by the UKB team, including reconstruction, gradient distortion correction, and quality control (QC). We briefly summarized the acquisition steps for each modality in Supplementary Text. Comprehensive information for the image acquisition and preprocessing is available at https://biobank.ctsu.ox.ac.uk/crystal/crystal/docs/brain_mri.pdf. We included 1,045 brain imaging traits in the study, including 230 sMRI traits for cortical and subcortical structures, 635 diffusion tensor imaging (DTI) traits from dMRI for white matter (WM) microstructure; and 90 rfMRI (tfMRI) traits for intrinsic (extrinsic) brain functions. For each trait and continuous covariate variable (discussed later), we removed values greater than five times the median absolute deviation from the median value.

We processed the sMRI locally using consistent procedures via advanced normalization tools (ANTs, http://stnava.github.io/ANTs). Specifically, we performed N4 bias correction, registration-based brain extraction, and a prior-based N4-Atropos 6 tissue segmentation (oasis template). The quality of segmentation was visually checked by experts. Subsequently, we conducted multi-atlas cortical parcellation based on the Desikan-Killiany-Tourville (DKT) cortical labeling protocol (*115*). After excluding three ROIs (5th ventricle, left lesion, right lesion) due to over 99% missing rate, 98 ROIs remained, with 62 of those capturing cortical regions. As a result, 101 traits for regional BVs were generated, including 98 regional and three global traits – gray matter volume (GMV), white matter volume (WMV), and total BV. The reproducibility, measured by the Pearson correlation between BV traits across two repeated scans from 2,944 retest subjects, was 0.92. For CT, we generated a global mean trait as well as regional traits from 62 cortical ROIs. Moreover, because the ANTs pipeline cannot generate surface area (SA) traits, we utilized 66 SA traits provided by the UKB (Category 193), which were derived based on the Desikan-Killiany (*116*) parcellation scheme using the FreeSurfer (https://surfer.nmr.mgh.harvard.edu) software. We provide details for the differences between DKT atlas and Desikan-Killiany atlas in Supplementary Text.

DTI evaluated dMRI in a tensor model and characterized water molecular diffusions in all directions. Five metrics of DTI: Axial diffusivity (AD), fractional anisotropy (FA), mean diffusivity (MD), mode of anisotropy (MO), and radial diffusivity (RD) were applied to each voxel of the image. The five DTI metrics can reflect different patterns of water diffusion in WM tracts. Specifically, AD is the eigenvalue of the principal direction; FA is related to directionality; MD quantifies the magnitude of absolute directionality; MO is the third moment of a tensor; and RD is the average of the eigenvalues of secondary diffusion directions. Given a metric, a tract-mean trait was generated by taking the average of all voxels in a tract. As a result, we obtained 110 traits, including 105 tract-mean traits for all tract-metric pairs and 5 overall-mean traits across all tracts ($21\times5 + 5 = 110$). We also applied functional principal component analysis (FPCA) to the voxels in a tract and extracted the top five functional PCs for each tract-metric pair. The PCs preserved the strongest variation in voxel-level measurement and are expected to provide more microstructural details on axonal organization and integrity. We generated 525 ($= 5\times5\times21$) functional PC traits for all tracts and metrics. The tracts were labeled by the ENIGMA-DTI pipeline (*117, 118*). Check Table S1 for the full names of the 21 tracts, and Supplementary Text for details of trait generation based on ENIGMA-DTI pipeline and FPCA.

The shape, face, and shape-face activation contrasts were the major interests in UKB for tfMRI. We extracted the time series from the whole scan of the tfMRI data including blocks of both shape and face activations (UKB Category 106). When we calculated the functional connectivity measurements in downstream analysis, we considered the shape and face contrasts together for capturing a more comprehensive view of the brain's functional dynamics.

We applied parcellation-based methods with Glasser360 (*38*) atlas (also called HCP-MMP) to generate 90 mean amplitude and functional connectivity traits each for rfMRI and tfMRI. The Glasser360 atlas is originally a surface-based parcellation for the cerebral cortex and has been transformed to a volumetric atlas to be compatible with the UKB volume-based data. We first projected the rfMRI and tfMRI data onto the Glasser360 atlas and generated 360×360 functional connectivity matrices. Then the 360 functional areas were grouped into 12 functional networks including the primary visual, secondary visual, auditory, somatomotor, cingulo-opercular, default mode, dorsal attention, frontoparietal, language, posterior multimodal, ventra multimodal, and orbito-affective networks (*119*). As a result, 12 mean amplitude traits and 78 (=12+11×12/2) mean pairwise functional connectivity traits were generated for the 12 functional networks. Refer to the Supplementary Text for introduction of fMRI acquisition and detailed steps of the parcellation-based dimension reduction procedure.

<u>Discovery and replication data processing for association analyses</u>

In our X-chromosome association analysis (XWAS), we utilized UK Biobank (UKB) phase 1-3 imaging data (up to February 2020) for the discovery phase, which included approximately 36,000 subjects. For replication, we employed phase 4 imaging data comprising an additional 3,100 subjects. We utilized the imputed genetic data (version 3). Details regarding genotyping and imputation are available in the UKB documentation. Genetic data processing in our study was conducted using PLINK2 (*52*) (v2.00a3LM, https://www.cog-genomics.org/plink/2.0/) separately for each imaging trait category.

In the discovery phase, we focused on subjects with non-Hispanic white ancestries (Field 21000). Following UKB's quality control guidelines, we excluded subjects with excessive heterozygosity (Field ID 22027), discrepancies between reported and genetic gender (Field ID 22001), potential sex chromosome anomalies (Field ID 22019), and a genotype missing rate over 5% (--mind 0.05). We excluded SNPs with an imputation score less than 0.3 (Resource 1967), a minor allele frequency (MAF) less than 0.0003 (--maf 0.0003) for heritability analysis (more on this later), and further excluded SNPs with an imputation score less than 0.6, a MAF less than 0.005 for XWAS. Moreover, SNPs with a Hardy-Weinberg equilibrium test p-value less than $10^{-6}$ (--hwe 0.000001) and multiallelic sites/indels (--snps-only just-acgt) were also excluded. To address potential relatedness among subjects, we employed GCTA (*42*) (v1.93.2 beta, https://yanglab.westlake.edu.cn/software/gcta/#Overview) to compute the genetic relationship matrix (GRM) using autosomes and excluded one of a pair of related subjects with the threshold 0.05 (--grm-cutoff 0.05), which removed approximately 1,800 subjects.

The refined discovery dataset consisted of 33,591subjects (15,939 males and 17,652 females) varying across imaging trait categories from 29,078 to 35,793. We analyzed 289,866 non-pseudo-autosomal region (NPR) SNPs and 11,508 pseudo-autosomal region (PAR) SNPs on the X-chromosome. For the replication phase, we included phase 4 non-Hispanic white subjects (UKBE, n = 4,181), phases 1-4 South Asian and Chinese subjects (UKBSAC, n = 462), and phases 1-4 African subjects (UKBA, n = 295). To enhance our sample size, we reintegrated white subjects previously excluded due to relatedness with the phase 4 white subjects (1,800 + 3,100), then conducted another round of relatedness pruning (--grm-cutoff 0.05). The same threshold was applied to Asian and African subjects for relatedness pruning. All other quality control steps for the replication phase were consistent with the discovery phase.

<u>Dosage compensation and heritability analysis</u>

We employed the GREML analysis tool in GCTA (*42*) for heritability analysis on NPR SNPs on the X-chromosome. We explored three model assumptions for GRMs on the X-chromosome: full DC (--dc 1), no DC (--dc 0), and equal variance (without specifying --dc) (*42*). These models account for differences in genetic variance between sexes due to their unique coding schemes. Specifically, females are consistently coded as (0, 1, 2), while males are coded as (0, 2) for full DC, (0, 1) for no DC, and (0, $\sqrt{2}$) for equal variance.

To identify the most suitable DC model for each brain imaging trait, we produced GRMs under each assumption. As default, we presumed a consistent allele frequency distribution between causal and genotyped SNPs and adjusted for imperfect LD (--grm-adj 0). Both GRMs for autosomes and the X-chromosome were included in a single model to jointly estimate heritability, enhancing the total heritability captured compared to separate analyses (*16*).

For sMRI and dMRI traits, we controlled for the indicator of release phase (1 if the subject was released in phase 3 and 0 if the subject was released in phase 1-2), assessment center (Field 54), genotype batch (Field 22000), top 40 genetic principal components (Field 22009), age at imaging, age-squared, sex, age-sex interaction, and age-squared-sex-interaction. Additional covariates were adjusted for non-global sMRI traits, including total BV, mean CT, and left/right total SA. For fMRI traits, following Alfaro-Almagro et al. (*120*), we additionally adjusted for head size (Field ID 25000), scan position X (Field ID 25756), scan position Y (Field ID 25757), scan position Z (Field ID 25758), scan table position (Field ID 25759), mean rfMRI head motion (Field ID 25741), and mean tfMRI head motion (Field ID 25742), as well as scan position X squared, scan position Z squared, mean rfMRI head motion squared and mean tfMRI head motion squared.

We employed the likelihood-ratio-test (LRT) with the null distribution $0.5\chi_1^2 + 0.5\chi_0^2$ to assess the X-chromosome's variance component in GREML. The Akaike information criterion (AIC) facilitated the comparison of model assumptions, with the smallest AIC indicating the optimal DC model.

<u>Enrichment analysis for heritability</u>

Following the approach of Mallard et al. (*16*), we characterized enrichment of X-linked heritability ($h_X^2$) by comparing it to the overall heritability ($h_{all}^2$) relative to the proportion of NPR variants on the X-chromosome. Variant counts for each chromosome were obtained from the Genome Reference Consortium Human Build 37 (GRCh37 release 13), available at https://www.ncbi.nlm.nih.gov/assembly/GCF_000001405.13. Specifically, the enrichment for $h_X^2$ is defined as $\frac{h_X^2/h_{all}^2}{length_x/length_{all}}$, where $length_x$ and $length_{all}$ represent the genomic lengths of the NPR on the X-chromosome and the entire genome, repectively. To determine whether $h_X^2$ was enriched or depleted for each trait, we applied a two-sided Z-test with the statistic $\frac{observed\ h_X^2 - expected\ h_X^2}{standard\ error\ of\ h_X^2}$, where $expected\ h_X^2 = \frac{h_{all}^2 \times length_x}{length_{all}}$, and the standard error of $h_X^2$ was derived by using GREML. For significant results, the Z-statistic greater than 0 indicates enrichment of $h_X^2$, while less than 0 suggests depletion.

<u>Sex-stratified heritability and phenotypic variance analysis</u>

We conducted a sex-stratified heritability analysis to directly examine the disparities in $h_X^2$ between males and females. Utilizing GREML, we incorporated the same set of covariates used in our sex-agnostic analysis, except for sex and its interaction terms. We adopted the equal variance model for all traits, which assumes that males and females have equivalent heritability. The GREML output provides the phenotypic variance, denoted as $V_p$. We subsequently calculated the phenotypic variance ratio for each trait as $\frac{V_{p,male}}{V_{p,female}}$.

<u>XWAS, sex-stratified XWAS, and meta-analysis</u>

We conducted association analysis for NPR and PAR SNPs using PLINK2 (v2.00a3LM). For NPR SNPs, we adopted the full DC model (--xchr-model 2) for traits that favor full DC or equal variance, and the no

DC model (--xchr-model 1) for others. The direction of effect size corresponded to the minor allele in the data. However, the minor allele for a particular variant might differ across imaging modalities. In the discovery phase, we adjusted for the same covariates as did in the heritability analysis. In the replication phase, the adjusted covariates included an indicator that the subject was included in phase 3 or phase 4, the assessment center, the top 10 genetic PCs, and all other imaging-related covariates. We applied the wild bootstrap approach to calibrate correlations between test statistics for imaging traits. The detailed procedure is provided in Supplementary Text. After multiple-comparison adjustment, SNPs achieving a genome-wide threshold of $5 \times 10^{-8}$ were considered as significant. Separate XWAS for males and females in the discovery cohort were conducted, maintaining consistent covariate adjustments, excluding sex and its interaction terms.

We employed FUMA (v1.4.1, https://fuma.ctglab.nl/) to group individual significant SNPs into genomic loci. The LD between SNPs were internally handled by FUMA using white subjects in 1000 Genome dataset. SNPs in LD > 0.6 were grouped under one independent significant SNP. Subsequently, those in LD > 0.1 were categorized under a single top SNP. LD blocks indexed by adjacent independent significant SNPs within 250 Kb of each other were merged into one genomic locus.

We employed a meta-analysis approach to integrate separate XWAS results using METAL (version released on 05.05.2020, https://genome.sph.umich.edu/wiki/METAL_Documentation). Inputs to the software included effect alleles, effect sizes, p-values, and sample sizes from individual analyses. The default procedure was adopted. Initially, p-values were transformed into Z-statistics. Then, the effect alleles and the direction of the effect sizes were adopted to align all studies to a consistent reference allele. An overarching Z-statistic was derived by taking a sample-size weighted sum of each individual statistic. The weighting was derived from the square root of sample size in each study. In our research, the meta-analysis combined the outcomes of sex-stratified XWAS, replication XWAS from UKBE, UKBSAC, and UKBA cohorts, as well as discovery and replication XWAS from UKBE participants.

Gene-level analysis and biological annotation

We conducted a series of gene-level analysis through FUMA (v1.4.1). We first performed a gene-based association analysis targeting 747 protein-coding genes on the X-chromosome using MAGMA (v1.08). In the analysis, SNPs were mapped to genes based on their physical locations in GRCh37, without including upstream and downstream regions of genes. Additionally, significant SNPs and other SNPs in LD > 0.6 (including those not in XWAS but from the 1000 Genome dataset) were passed to functional consequence mapping (ANNOVAR (*121*), version 2017-01- 11), eQTL mapping (with reference database: CommonMind Consortium (*20*) and GTEx v8 brain), and 3D chromatin interaction mapping (built-in chromatin interaction data: adult cortex, fetal cortex, dorsolateral prefrontal cortex, and hippocampus (*122*); annotate enhancer/promoter regions: E053-E082 (brain)).

All significant genes prioritized in the functional mapping analyses were annotated using the DAVID Bioinformatics Database (*94*) (https://david.ncifcrf.gov/home.jsp) and SynGO (https://syngoportal.org/), with all parameters retained as default.

We further employed H-MAGMA (*123*) to map significant SNPs to long non-coding RNAs using chromatin interaction profiles. An annotation file specific to the X-chromosome was created following established protocols (*124*) and applied MAGMA (v1.08) for H-MAGMA execution. Minor adjustments were made to the R code to focus exclusively on the X-chromosome.

Summary data-based Mendelian randomization (SMR)

The summary statistics of the XWAS and the summary statistics of eQTL analysis using CAGE whole-blood data provided by Sidorenko et al. (*44*) were utilized in the SMR analysis. The genetic data in XWAS was used as a reference for LD estimation. There were 1,639 probes for genes in the NPR of the

X-chromosome, and significant trait-gene pairs were identified by controlling FDR at 0.05 level. The significance indicates that the gene expression level may have a causal effect on the trait. Then we performed the HEterogeneity In Dependent Instrument (HEIDI) test to distinguish the pleiotropy of causal SNPs from linkage for the significant trait-gene pairs. A non-significant result at the nominal level (p-value > 0.05) corresponds to no linkage effect, thereby supporting the hypothesis of pleiotropy.

Sex differences in genetic effect

Sex differences in genetic effects of NPR SNPs were tested by a two-sided z-test (*95*). The null hypothesis is $H_0$: $\beta_m = \beta_f$, where $\beta_m$ and $\beta_f$ are true per-allele genetic effects for males and females, respectively. We coded males as (0, 2) for a full DC trait, and as (0, 1) for a no DC trait while always coded females as (0, 1, 2). The test statistic is defined as $z = \frac{b_m - b_f}{\sqrt{se_m^2 + se_f^2}}$, regardless of DC assumption, where $b$ is a genetic effect estimate and $se$ is the corresponding standard error. The raw SNP effect size and its standard error were divided by the trait's standard deviation derived from the phenotypic variance to correct for scale effects that could act as confounders in the study.

Differences in genetic profiles between subjects classified by phenotypic quantiles

We initially segregated the data by sex, considering significant confounding effects from sex-related phenotypic differences. For every trait, we calculated both the upper and lower 10th percentile scores. Using these scores, we selected subjects based on their quantile rankings. For instance, within a specific trait category (e.g., BV), if a subject's scores for over 25% of the traits surpassed the upper 10th percentile for those traits, then the subject was categorized into the "upper outlier" group. Conversely, if a subject's scores for more than 25% of the traits fell below the lower 10th percentile, then the subject was placed in the "lower outlier" group. We then extracted the genetic profiles of these subjects using PLINK2 (--geno-counts). Male genetic profiles were coded as 0 or 2, while female profiles were coded as 0, 1, or 2. We employed Fisher's exact test to compare genetic profiles between the "upper outlier" and "lower outlier" groups for each sex.

**Supplementary Text**

An introduction to imaging acquisition in the UK Biobank

The acquisition of T1-weighted structural scans was conducted using Siemens Skyra 3T scanners, employing a 3D MPRAGE sequence to achieve high-resolution imaging. These scans featured 1-mm sagittal slices and a 1×1-mm in-plane resolution, with in-plane acceleration (iPAT) set to 2, alongside prescan normalization to ensure consistent image quality across the dataset. To maintain the confidentiality and anonymity of study participants, the T1 images underwent a defacing process, removing any facial features that could potentially identify individuals.

The UKB dMRI data collection protocol was designed to capture high-resolution brain microstructures, utilizing a 2×2×2 mm spatial resolution with anterior-to-posterior phase-encoding and a multiband acceleration factor of three. The imaging employed two nonzero b-values of 1000 and 2000 s/mm$^2$, each with 50 diffusion-encoding directions to enhance the detection of microstructural properties of the brain's WM. The diffusion preparation followed a standard Stejskal-Tanner pulse sequence, characterized by an echo time (TE) of 92 ms and a repetition time (TR) of 3600 ms. This choice aimed to improve the signal-to-noise ratio (SNR) by opting for a shorter TE compared to a twice-refocused (bipolar) sequence, albeit with a trade-off of increased susceptibility to eddy current distortions. For a more detailed description of the dMRI data acquisition protocol and its specifications, refer to Section 2.8 of the UKB brain imaging documentation.

The UKB rfMRI and tfMRI imaging data were collected using Siemens Skyra 3T scanners, following standardized protocols. For rfMRI, data was captured over 490 time points during a six-minute span, with a spatial resolution of 2.4×2.4×2.4 mm and dimensions of 88×88×64. The TE was set at 39 ms, and the TR at 735 ms. Additionally, a single-band reference scan, matching the time series data's geometry but with enhanced between-tissue contrast, was acquired for head motion correction and modality alignment purposes (*125*).

The tfMRI data utilized the Hariri faces/shapes emotion task, adapted from the Human Connectome Project (HCP) with modifications for duration and stimulus block repeats. (https://biobank.ndph.ox.ac.uk/showcase/label.cgi?id=106). Participants engaged with blocks of trials, identifying matching faces or shapes. This task was conducted over 332 time points within four minutes, maintaining the same spatial resolution, dimensions, and scanning parameters as the rfMRI sessions. The shape, face, and shape-face activation contrasts were the major interests in UKB.

Preprocessing of both rfMRI and tfMRI datasets was conducted by the UKB brain imaging team (*125*). The full pipeline can be found in Section 3 of the UKB Brain Imaging Documentation. Overall, the pipeline encompassed image cleaning, image registration, and representative time series generation. The detailed preprocessing pipeline is documented by the UKB, with source codes accessible at https://git.fmrib.ox.ac.uk/falmagro/UK_biobank_pipeline_v_1.

<u>The differences between DKT atlas and Desikan-Killiany atlas in brain anatomy</u>

In general, the DKT atlas is very similar to Desikan-Killiany atlas as the former parcellates each hemisphere into 31 regions while the latter parcellates it into 32. The 31 cortical labels in the DKT atlas are actually a modification of the Desikan-Killiany atlas to improve cortical labeling consistency and enhance FreeSurfer's cortical classification. Specifically, the DKT atlas excluded the regions "banksst" and "frontal pole" but included the "insula" region. All other ROIs exactly match in two atlases.

<u>The ENIGMA-DTI pipeline and functional principal component analysis for DTI</u>

Our analysis employed the ENIGMA-DTI pipeline, a standardized set of procedures for processing DTI data across various datasets (http://enigma.ini.usc.edu/protocols/dti-protocols/). The process began with linear registration, aligning each FA image to the ENIGMA FA template, which is set at a 1×1×1 mm spatial resolution in the MNI-ICBM-152 standard space. We then applied nonlinear registration techniques to further refine the alignment of the FA images to the standard space. The registered FA images were then masked using the template brain mask to isolate brain tissue from non-brain elements within the images. Subsequently, we transferred the individual images of AD, MD, MO, and RD into the FA template space. The ENIGMA skeleton, representing the major WM pathways, was projected onto the registered images. Finally, we extracted tract-based statistics for each DTI metric, including tract-averaged means and functional PCs.

To generate PCs for DTI metrics, we employed a "smooth-first-then-estimate" method, detailed at https://doi.org/10.5281/zenodo.4549465, using FA as an example. Initially, we extracted skeletonized FA images from 17,706 subjects in UKB phases 1 and 2, smoothing them with FSL software using a 2 mm Gaussian kernel. Smoothed voxel-wise FA values for each tract were then centralized by subtracting the dataset's group average. Subsequently, these values were reshaped into an $n \times p$ matrix A, where n is the sample size and p is the size of the tract. We performed singular value decomposition $A = UDV^T$, where V is a $p \times p$ orthogonal matrix, U is an $n \times n$ orthogonal matrix, and D is a diagonal matrix. The top 5 FA PCs were extracted, explaining over 70% of the FA variation on average.

<u>Parcellation-based network level feature extraction using the Glasser360 atlas</u>

The Glasser360 atlas(*38*), a comprehensive parcellation developed from high-resolution functional and anatomical MRI data from Human Connectome Project (HCP) participants, offers a detailed representation of the cortex with 360 distinct cortical areas (180 per hemisphere). For our analysis, we utilized the volumetric version of the Glasser360 atlas, available at NeuroVault (https://identifiers.org/neurovault.collection:1549), which had been transformed from its original surface-based format into a volumetric atlas via FreeSurfer. This conversion allows for direct application to the rfMRI and tfMRI datasets subject to appropriate preprocessing.

The Glasser360 atlas was applied to these datasets to extract regional time series for all 360 cortical areas. From these time series, we derived amplitude traits based on standard deviation and full connectivity matrices through Gaussianized temporal correlations between all pairs of node time series. Further, by adopting the network definitions from Ji et al. (*119*), which split the 360 areas into 12 networks, we generated network-level traits. This segmentation resulted in 78 network-level groups, encompassing 12 within-network and 66 between-network groupings.

Adjusting for multiple hypothesis testing in XWAS by wild bootstrap

Suppose we have $Q$ traits in total, and we want to adjust for multiple hypothesis testing considering the number of traits by using wild bootstrap approach, which indicates that we still apply the genome-wide threshold $5 \times 10^{-8}$ for significance to the adjusted p-values. The specific steps are as follows,

1. For each trait $Y_q$, fit a null model $Y_q = X_q\alpha_q + \varepsilon_q$, where $Y_q$ is a vector of trait; $X_q$ is the covariate matrix for trait $q$. We get $\hat{\alpha}_q$ and $\hat{\varepsilon}_q$.
2. Let $v_i^{(b)} \sim N(0,1)$, generate a sample $b$ for each trait and each subject $i$: $Y_{iq}^{(b)} = X_{iq}'\hat{\alpha}_q + v_i^{(b)}\hat{\varepsilon}_{iq}$. Note that $v_i^{(b)}$ are the same for all traits but different among subjects and bootstrap samples.
3. For a SNP $k$, $k = 1, \dots, N_g$, do XWAS on all traits with models $Y_q^{(b)} = X_q\alpha_q^{(b)} + Z_{qk}\beta_{qk}^{(b)} + \varepsilon_q^{(b)}$, $q = 1, \dots, Q$, where $Z_{qk}$ are genotype vectors with different sample sizes across traits. Then we get $F$ statistics for all $Q$ traits $(F_{1k}^{(b)}, \dots, F_{Qk}^{(b)})$'. Take the max $F$ statistic $F_k^{(b)*}$ over all traits.
4. Repeat the step 2, 3 for $B = 150$ samples, we get $(F_k^{(1)*}, \dots, F_k^{(B)*})$'. Then we approximate a $\chi^2$ distribution for SNP $k$ by moment matching. Specifically, let $k_1$, $k_2$ and $k_3$ denote the mean, variance, and skewness of $(F_k^{(1)*}, \dots, F_k^{(B)*})$'. We compute parameters $a = k_3/(4 \times k_2)$, $b = k_1 - 2k_2^2/k_3$, and $d = 8k_2^3 / k_3^2$. The adjusted p-value for SNP $k$ on trait $q$ can be approximated by $1 - pchisq(\frac{F_{qk}^{obs} - b}{a}, d)$ , where $pchisq$ is the c.d.f. of $\chi^2$ distribution, and $F_{qk}^{obs}$ is the observed F statistic computed from the real data. Note the more bootstrap samples, the more accurate the approximation. If the raw p-values are extremely small, such as $10^{-20}$, $B$ may increase to 200 or 300.

Since the computational burden of implementing the procedure for all SNPs was intractable, we split the SNPs into five groups based on MAF: $[0.005, 0.01)$, $[0.01, 0.05)$, $[0.05, 0.10)$, $[0.10, 0.25)$, and $[0.25, 0.50]$, and randomly picked 10 SNPs in each group. Within a group, we combined $(F_k^{(1)*}, \dots, F_k^{(B)*})$ generated from the 10 SNPs to approximate the $\chi^2$ distribution for that group. That implies we implicitly assumed that the distribution parameters were identical for SNPs in the same MAF group.

Methods for sex phenotypic difference analysis

We used linear models including sex and other covariates used in XWAS to test sex differences. Sex was the term of interest (males coded as 1 and females coded as 2) and the significance was determined via an F test. A negative effect size of sex indicated males had greater measurement than females. Note that we also included the interaction terms between sex and age (and age squared) in the model, but we only focused on the main effect of sex in assessment.

Additional results on dosage compensation and X-linked heritability for DTI and fMRI traits

Out of the 110 tract-mean traits, 93 exhibited significant $h_X^2$, with an average $h_X^2$ of 1.2% (se = 0.39%) (Fig. S16), whereas 330 out of 525 DTI PC traits were significant, with an average $h_X^2$ of 1.3% (se = 0.42%) (Fig. S17). The atlas of enrichment for tract-mean DTI traits are present in Fig. S18.

For each pair of tract and DTI metric, there was a correspondence between the number of PCs favoring full DC and the DC status of the tract-mean trait (Fig. S19). Most tract-mean traits in favor of full DC had more than three PCs also favoring full DC. In contrast, those favoring no DC typically had no more than two PCs in agreement. However, we found some outliers. For example, FA of the fornix had the second to fifth PC favoring no DC, but the tract-mean trait favored full DC. Similarly, MO of the posterior corona radiata had the second to fifth PC favoring full DC but the tract-mean trait favored no DC. The latter case is probably because the first PC had extremely high $h_X^2$ of 4.6%, thus dominating the DC status. As a result, the top five PCs can capture the major genetic information underlying the DC mechanism for DTI traits. The heritability analysis for DTI traits on the X-chromosome shows that functional PCs can provide different dimensions from the tract-mean traits to explain genetic control.

Distinct patterns in $h_X^2$ emerged between rfMRI and tfMRI traits. For rfMRI, language, secondary visual, and frontoparietal networks had the highest $h_X^2$ among the mean amplitude traits ($h_X^2 = 1.67\%{\sim}1.79\%$), while the average functional connectivity within primary and secondary visual networks and the one between these two networks surpassed all other connectivity traits ($h_X^2 = 2.46\%{\sim}2.86\%$) (Fig. S20A). However, tfMRI traits presented a different hierarchy of heritability. The mean amplitudes of frontoparietal, posterior-multimodal, and dorsal-attention networks were the most heritable ($h_X^2 = 1.13\%{\sim}1.53\%$), while the average functional connectivity traits between frontoparietal and posterior-multimodal networks, and that between language and posterior multimodal networks, as well as that within the language network surpassed all other tfMRI triats ($h_X^2 = 1.78\%{\sim}1.94\%$) (Fig. S20B).

<u>Gene-based analysis and Functional gene mapping</u>

MAGMA (*126*) (v1.08) was applied to XWAS summary statistics for gene-based analysis, which aggregated individual variant effects to evaluate association signals of genes. We identified 45 genes in NPR located at 23 genomic regions, associated with 137 traits by controlling for FDR at 0.05 level (Table S20). *CLIC2*, *DUSP9*, *RAB39B*, *TMLHE*, *VBP1*, and *PJA1* were previously seen as being associated with regional SA phenotypes (*16*). *ZNF275*, *DACH2*, and *VMA21* were associated with brain connectivity measurement (*127*) and brain shape (*55*). *FAAH2* was associated with neuroticism measurement (*18*). Many detected genes were involved in intellectual disability, educational attainment, and neuropsychiatric disorders. For instance, *RENBP* (*128*), *TKTL1* (*58*), and *MAP7D2* (*58*) were for schizophrenia; *USP51* was for educational attainment (*5, 60*); *DACH2* was for AD (*129*). *IRAK1* and *TMEM187* were for Internet addiction disorder (*130*). Some genes were linked to subtypes of autism. For instance, *TMLHE* deficiency, leading to a defect in carnitine biosynthesis, was a risk factor for nondysmorphic autism (*91, 92*). *DACH2* was associated with ASD (*131*).

We mapped the significant SNPs to genes based on functional sequences, resulting in 38 unique genes associated with 72 traits. These genes exhibited diverse association patterns with SNPs that influenced brain measurements, neuropsychiatric disorders as well as cognitive abilities (Table S21). For instance, *DCAF8L1* was linked to educational attainment (*60*) and neuroticism (*18*), *EFNB1* was linked to schizophrenia (*61*), Parkinson's disease (*64*) and educational attainment(*60*), while both *RN7SKP31* and *RNU6-985P* were linked to neuroticism (*132*), cognitive function (*133*) and educational attainment (*60*), through SNPs in nearby intergenic regions. Moreover, *SRPX* was tagged by rs35318931 in its exonic region, correlating with CSF volume, which was a risk factor for anorexia nervosa (*134*).

X-chromosome inactivation (XCI) status (*6*) was determined for all protein-coding genes identified by FUMA (Table S22). Among the 96 genes with available XCI information, 65 (67.7%) were previously reported inactive, 12 (12.5%) were escaping, and 19 (19.8%) were variable. When compared with the reference provided by Tukiainen et al (*6*), the identified genes were not enriched in any XCI category (hypergeometric test, p-value > 0.05/3).

Long non-coding RNAs (lncRNAs) have merged pivotal regulators. For example, *XIST* and *TIST* not only initiate XCI but also participate in subsequent complex processes (*135, 136*). We utilized two approaches – eQTL mapping and H-MAGMA (*123*) to map significant SNPs to lncRNAs. H-MAGMA incorporates a chromatin interaction profile to aggregate SNPs to the nearest genes (Methods, Table S23). SNPs associated with 82 traits were mapped to 14 lncRNAs across 10 genomic regions by controlling FDR at the 0.05 level.

Sex phenotypic differences in complex brain imaging traits

Sex differences in the human brain with respect to cortical and subcortical structures, WM microstructures, and functional connectivity were observed in plenty of studies (*97*). We evaluated the sex differences using a linear model and adjusted for potential confounding factors that may bias the sex effect, including age, genetic principal components, head motion and positions, etc (Supplementary Text). Out of the total 1,045 traits, 920 (88.0%) were significantly different between sexes after controlling for FDR at 0.05 level (Table S14). Notably, each category of brain imaging traits maintained a comparable proportion of significant differences. We then focused on these significant traits in the following analysis.

Set males as reference (males were coded as 1, females were coded as 2), a negative effect size of sex means males had a greater measurement. The mean effect size of the significant traits was -0.05, indicating males registered a 5% standard deviation increase compared to females. And 60.5% significant traits had a negative effect size for sex (proportion test against 0.5, p-value = 0.03). The proportion of negative effect size varied across trait categories (Fig. S21). For instance, both CT and BV had more than 58% traits with negative effects for sex, but the proportion for SA was only 47.4%. All DTI traits exhibited the proportion greater than 50%, where 71.8% tract-mean traits exhibited larger values for males. Remarkably, 79.3% rfMRI traits were larger for males, while the proportion for tfMRI traits was decreased to 67.8%. In parallel with the proportion results, tract-mean DTI traits and rfMRI traits displayed the strongest sex bias with the mean effect size less than -0.1.

By closely examining the sex differences within each modality, we found more significant disparities. For example, males had larger total BV, cerebrospinal fluid volume, and total SA relative to females, but had smaller gray and white matter volume and mean CT. Overall DTI mean traits were larger for males except for RD. The external capsule and the ratio of fornix to stria terminalis were the only two tracts with negative sex effect evaluated by AD. For rfMRI and tfMRI traits, we discovered that males had greater mean amplitude for all networks, but females had greater mean functional connectivity related to the ventral-multimodal network. Most of these discoveries were consistent with previous studies (*30, 137-141*).

Interestingly, we found significant negative correlation between the sex differences and $h_X^2$ (r = -0.17, p-value = $3.8 \times 10^{-7}$), which means traits with larger measurements in males tend to have greater $h_X^2$. We speculate that this correlation may be explained by larger phenotypic variance in males. We also observed significant correlation between male:female variance ratio and $h_X^2$ (r = 0.17, p-value = $1.6 \times 10^{-7}$). Estimation of the phenotypic variance ratio is provided in Methods and the results can be found in Table S2. However, there was no significant signals between sex phenotypic differences and variance ratio (r = -0.03, p-value = 0.38). We found no significant disparities in sex phenotypic differences among the DC status (Wilcoxon rank sum test, p-value > 0.19), nor did we find significant disparities in sex phenotypic differences among $h_X^2$ enrichment groups (Wilcoxon rank sum test, p-value > 0.22).

There is increasing evidence that both the X-chromosome and sex steroid exposures across the lifespan can potentially alter the human brain from many aspects (*4, 8, 32, 142*), and therefore, we are motivated to investigate genetic underpinnings of human brain with respect to the X-chromosome.

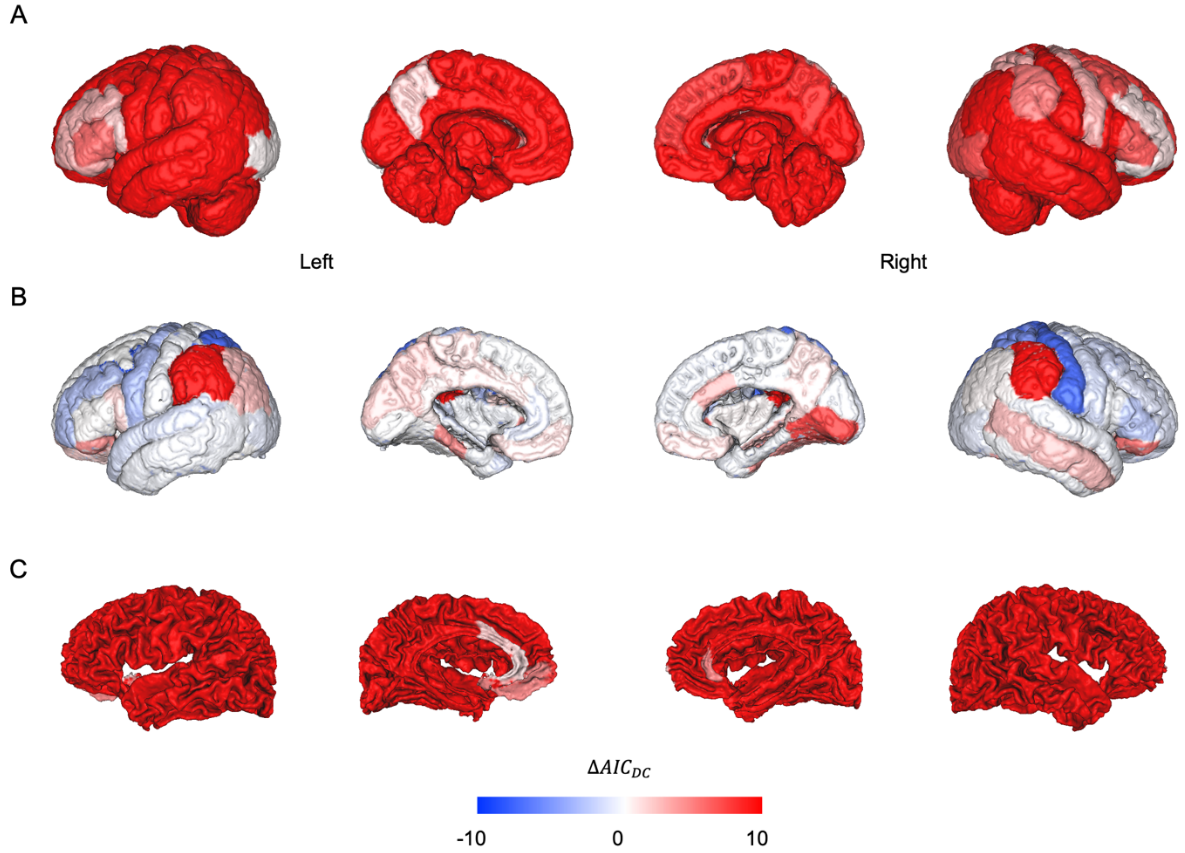


**Fig. S1: Continuous map showing regional variation in the strength of statistical evidence for X-chromosome dosage compensation in sMRI traits, indexed by $\Delta AIC_{DC}$.** There is evidence for full DC if $\Delta AIC_{DC}$ is greater than zero, while it indicates no DC if $\Delta AIC_{DC}$ is less than zero. It equals zero when the equal variance model fits best. Any $\Delta AIC_{DC}$ that are greater than 10 or less than -10 are truncated to 10 and -10, respectively, for illustration. A) BV; B) CT; and C) SA.

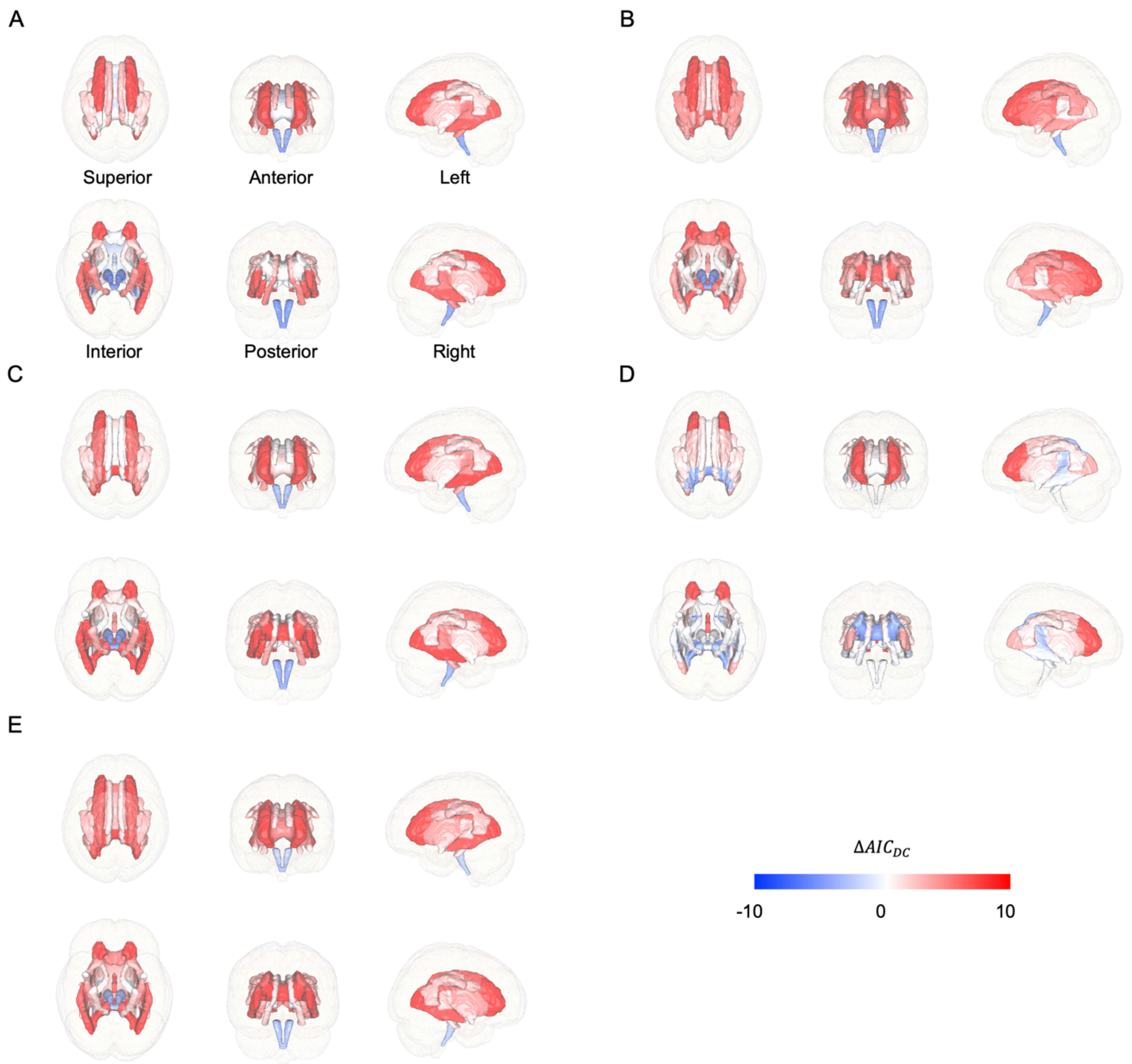


**Fig. S2: Continuous map showing regional variation in the strength of statistical evidence for X-chromosome dosage compensation in DTI traits, indexed by $\Delta AIC_{DC}$.** There is evidence for full DC if $\Delta AIC_{DC}$ is greater than zero, while it indicates no DC if $\Delta AIC_{DC}$ is less than zero. It equals to zero when the equal variance model fits best. Any $\Delta AIC_{DC}$ that are greater than 10 or less than -10 are truncated to 10 and -10, respectively, for illustration. A) AD; B) FA; C) MD; D) MO; and E) RD.

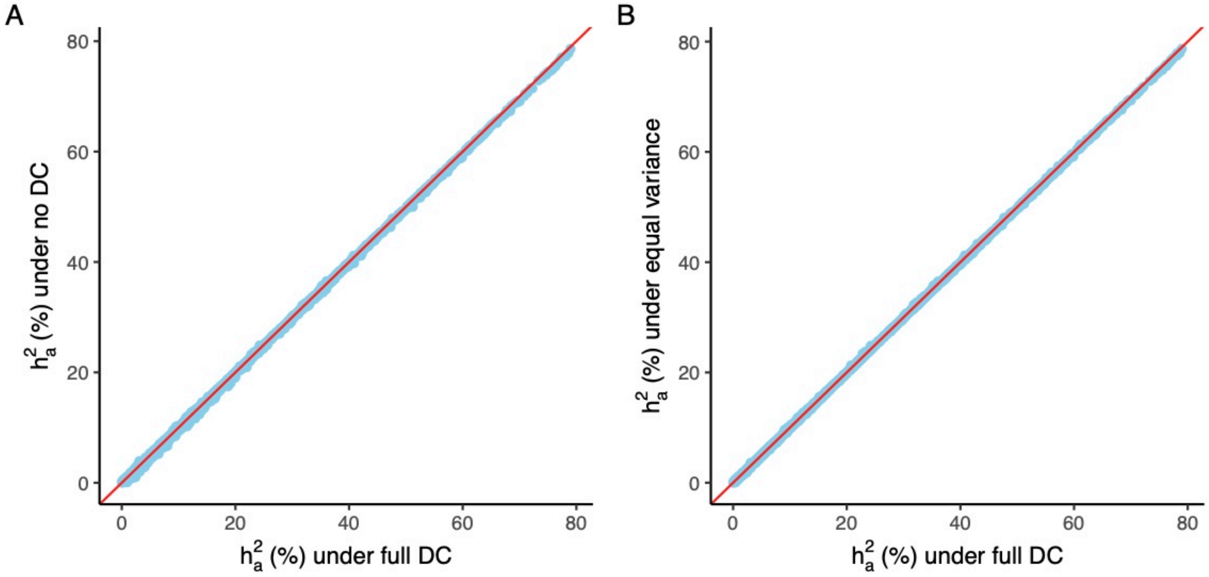


**Fig. S3: Comparisons of autosomal heritability ($h_a^2$) among DC groups.** The $h_a^2$ is estimated by the model including both GRMs of the autosomes and the X-chromosome in GCTA. A) Comparison between using full DC and no DC. B) Comparison between using full DC and equal variance.

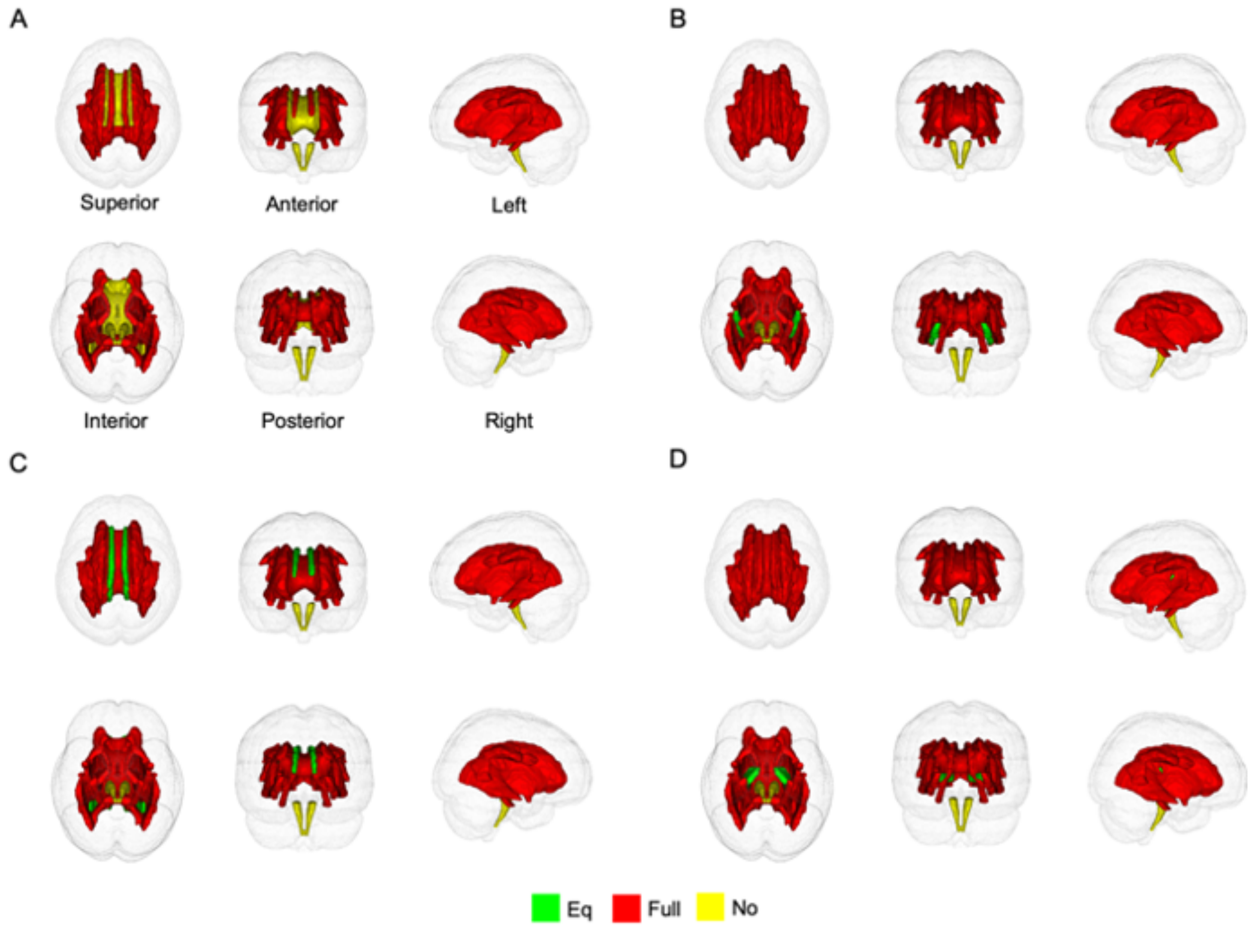


**Fig. S4: Atlases of DC in DTI tract-mean traits evaluated by different metrics.** A) AD; B) FA; C) MD; and D) RD.

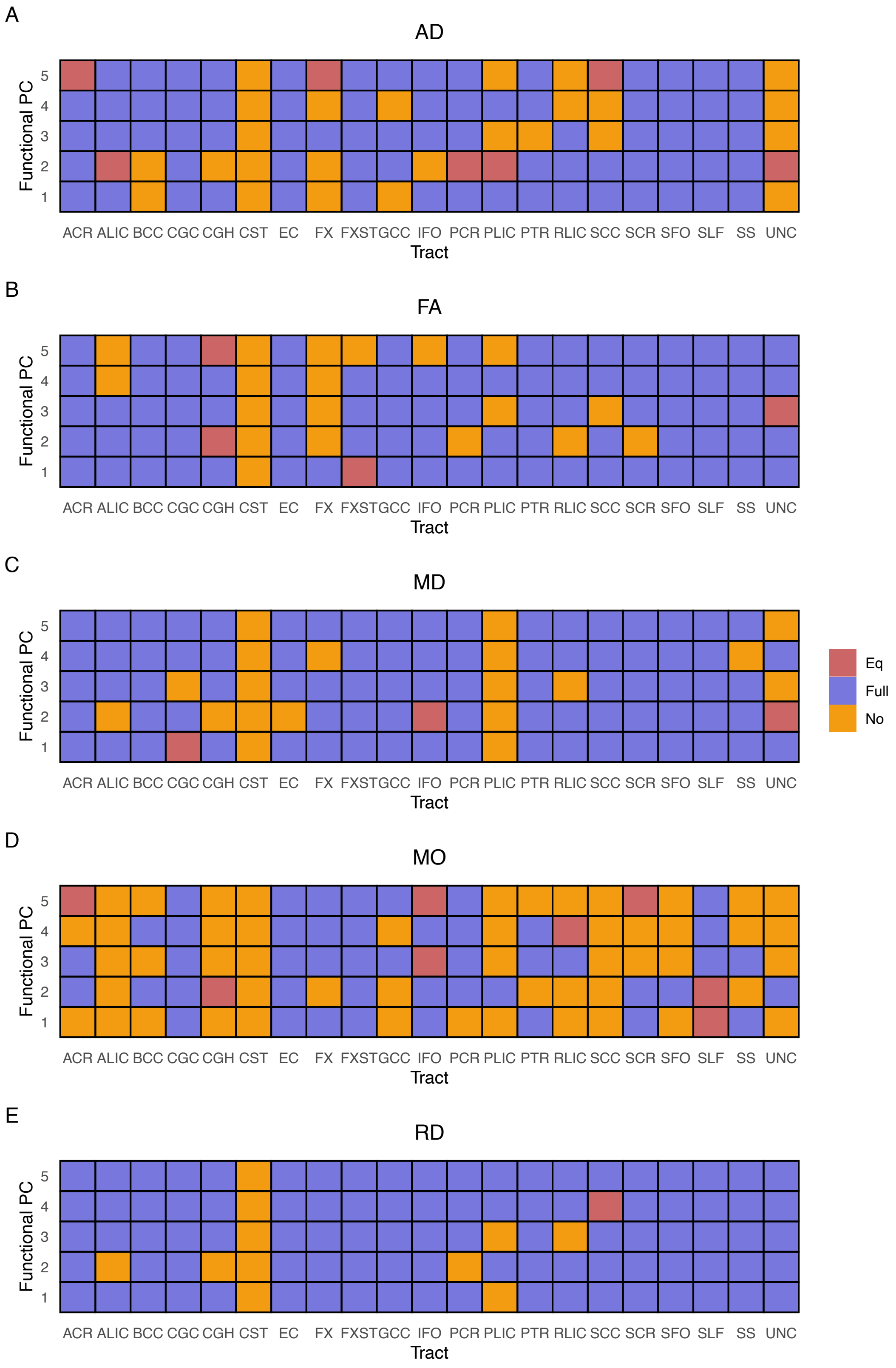


**Fig. S5: Atlases of DC of DTI PC traits evaluated by different metrics.** A) AD; B) FA; C) MD; D) MO; and E) RD.

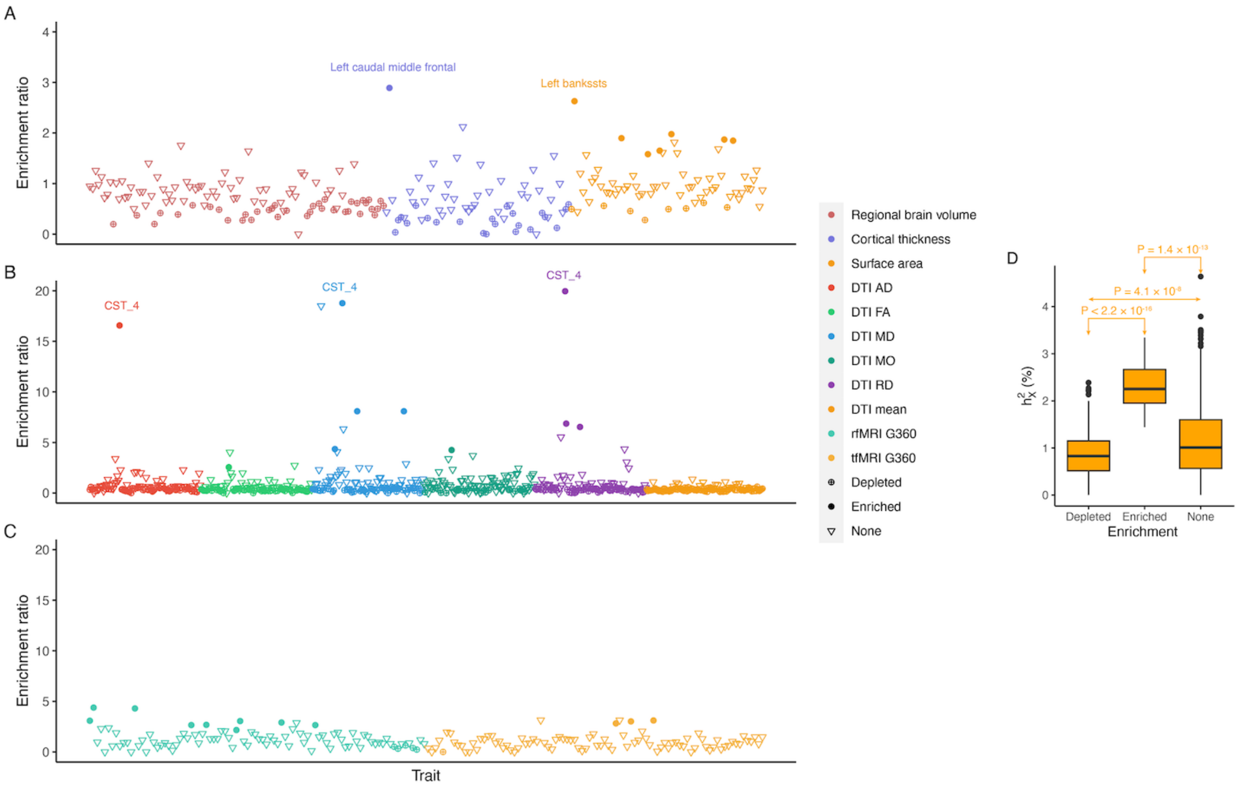


**Fig. S6: Enrichment analysis of X-linked heritability ($h_X^2$) for complex brain imaging traits.** A)-C) Scatter plots of enrichment ratios for individual traits in sMRI, DTI, and fMRI, respectively. The different trait categories are distinguished by color. Traits with significant enrichment are shown in solid circles, while traits with significant depletion are shown in crossed circles. Traits with no significant signals are shown in inverted triangles. Some traits with exceptionally high enrichment ratios are annotated. "CST_4" means the fourth PC of corticospinal tract evaluated by a particular metric represented by color. D) Pairwise comparisons of $h_X^2$ estimates between enrichment groups. The p-values are the results of the Wilcoxon rank sum test.

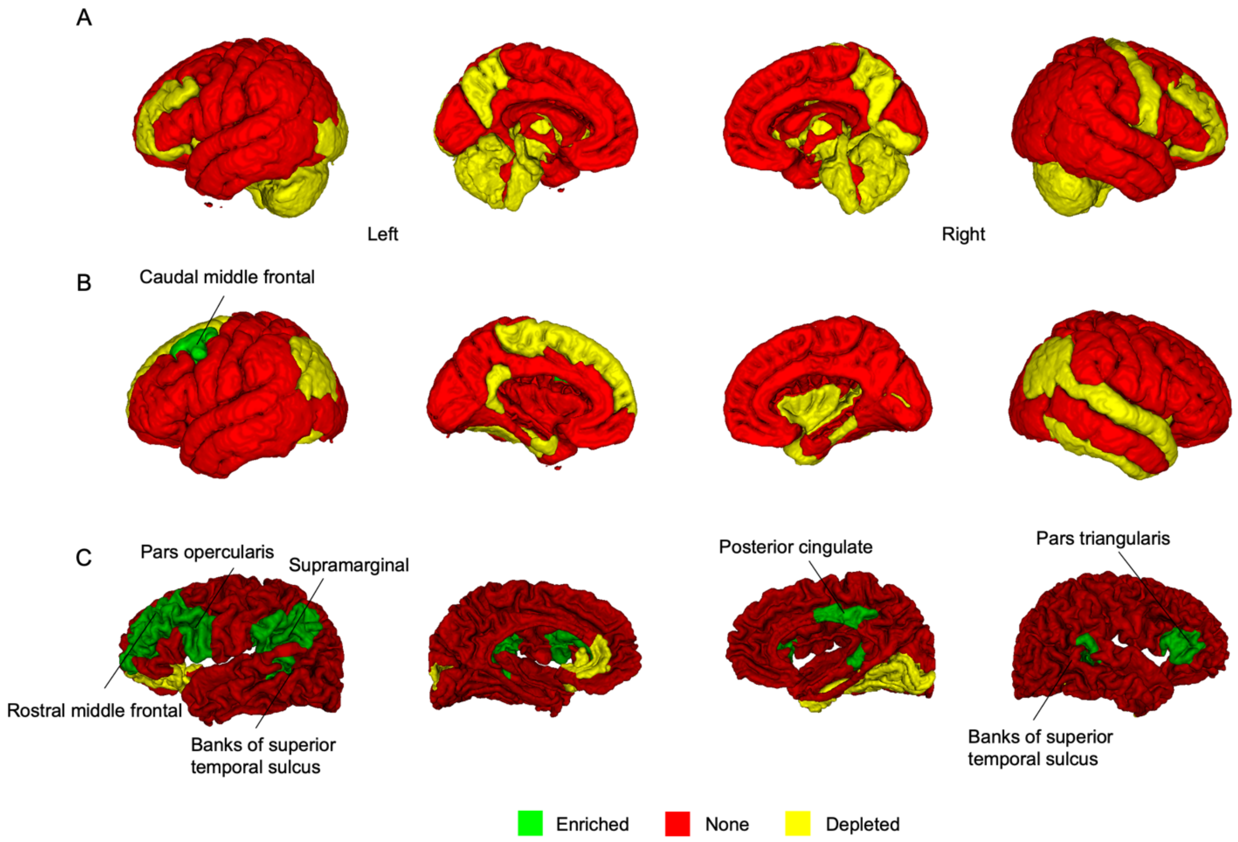


**Fig. S7: Regional patterning of X-linked heritability enrichment for sMRI traits.** Significantly enriched traits are annotated. A) regional BV; B) CT; and C) SA.

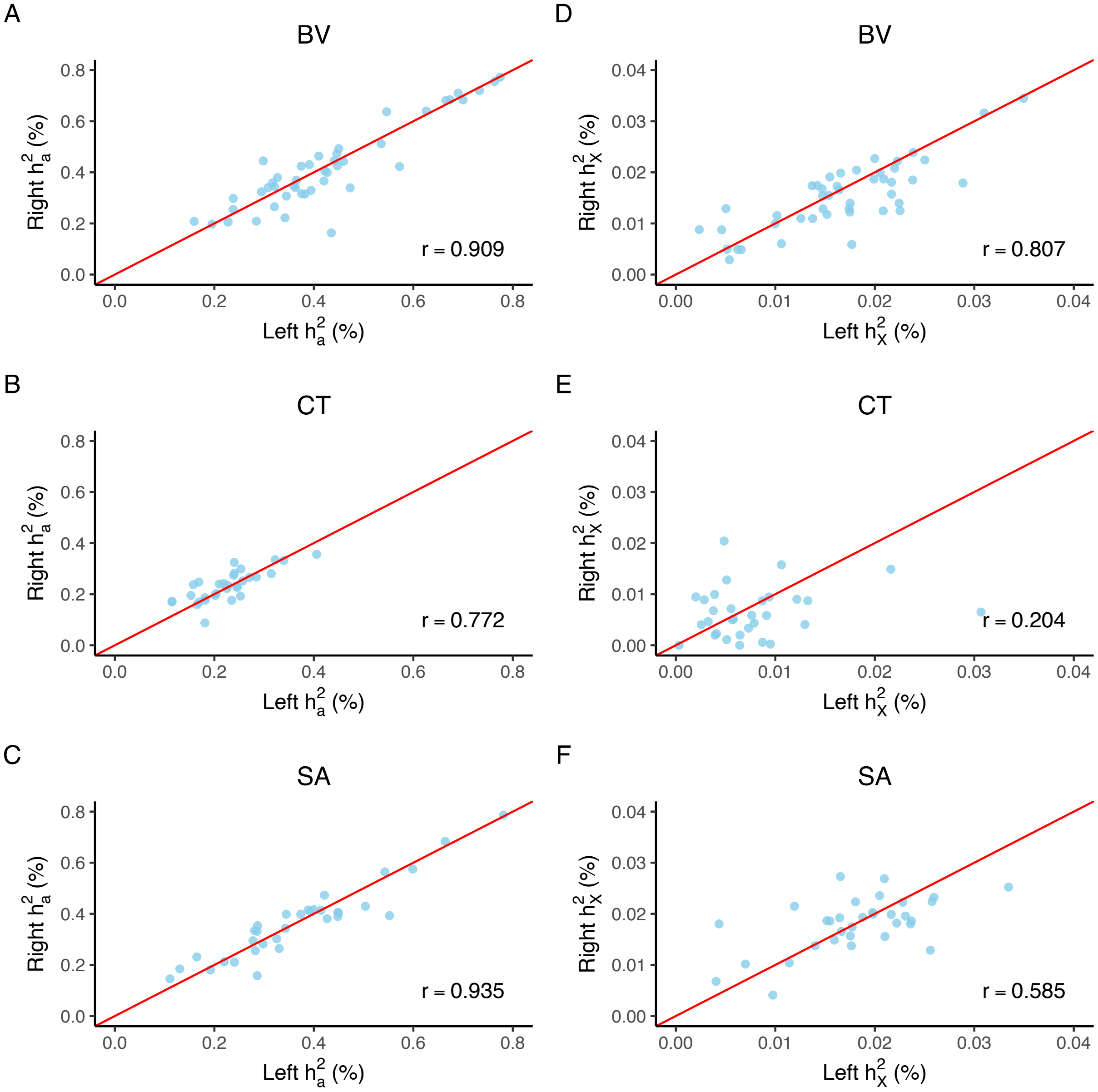


**Fig. S8: Symmetricity of X-linked heritability between left and right hemispheres for brain anatomy.** The Pearson correlation coefficient is annotated on each plot. A)-C) Autosomal heritability ($h_a^2$). D)-F) X-linked heritability ($h_X^2$). Only the coefficient in panel E is not significant at 0.05 level (p-value = 0.20).

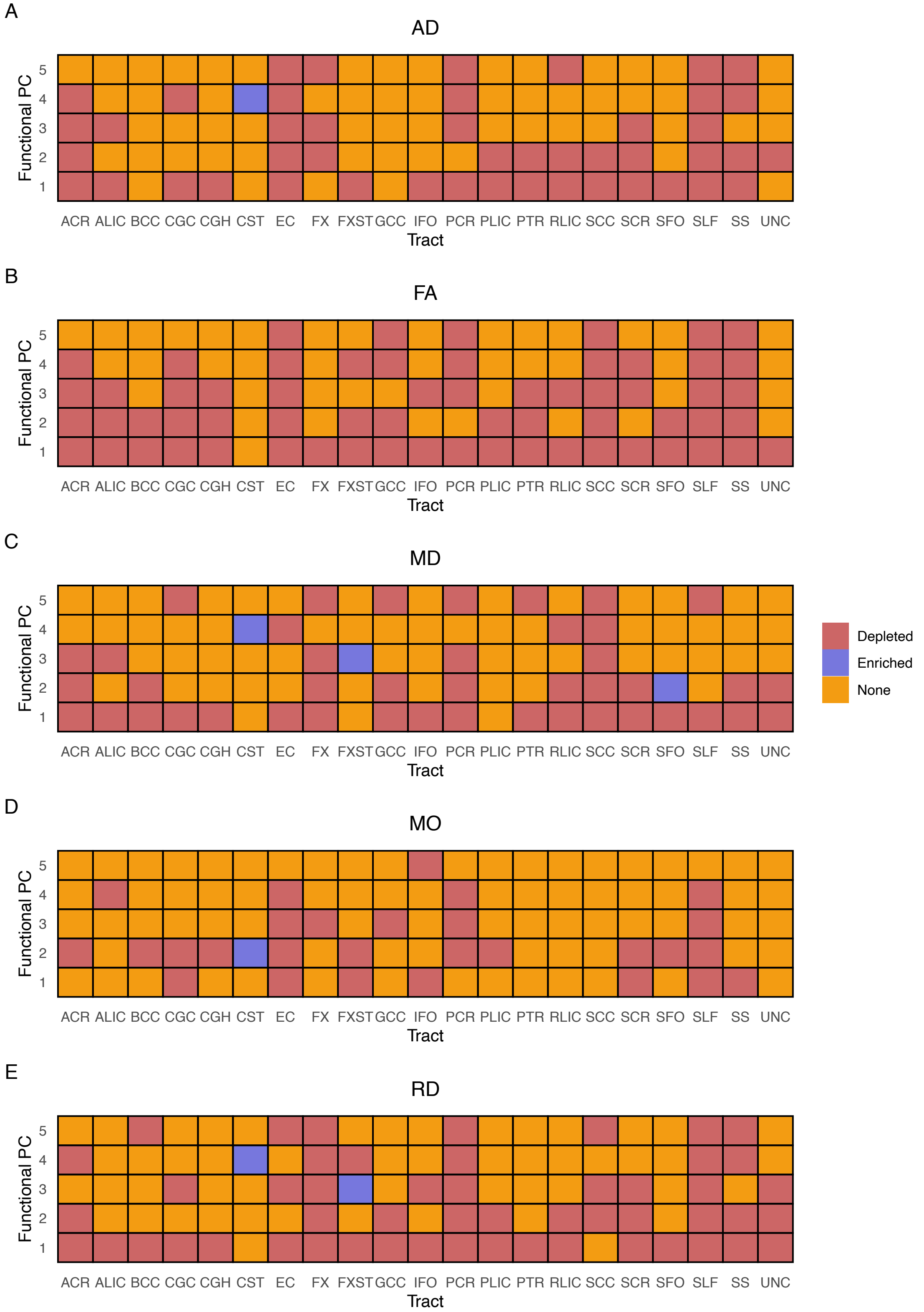


**Fig. S9: Atlases of enrichment of DTI PC traits evaluated by different metrics.** A) AD; B) FA; C) MD; D) MO; and E) RD.

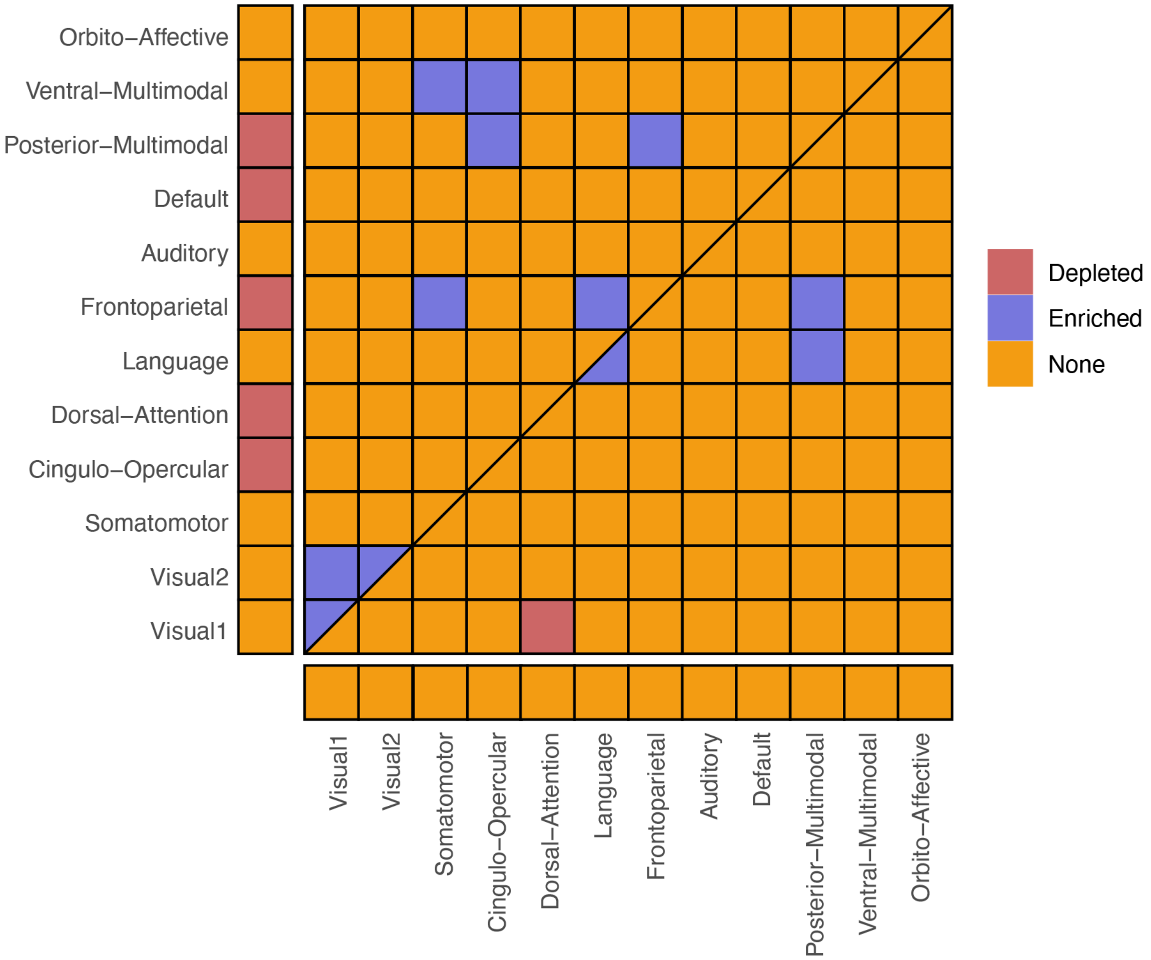


**Fig. S10: An atlas of enrichment for fMRI traits.** The upper triangle represents rfMRI, while the lower triangle represents tfMRI. The diagonal extending from the bottom left to the top right, is split into two triangles: the upper one represents enrichment for intra-connectivity of rfMRI and the lower for tfMRI. Cells on the left margin represent enrichment for amplitude traits for rfMRI, while those at the bottom indicate enrichment for amplitude traits for tfMRI.

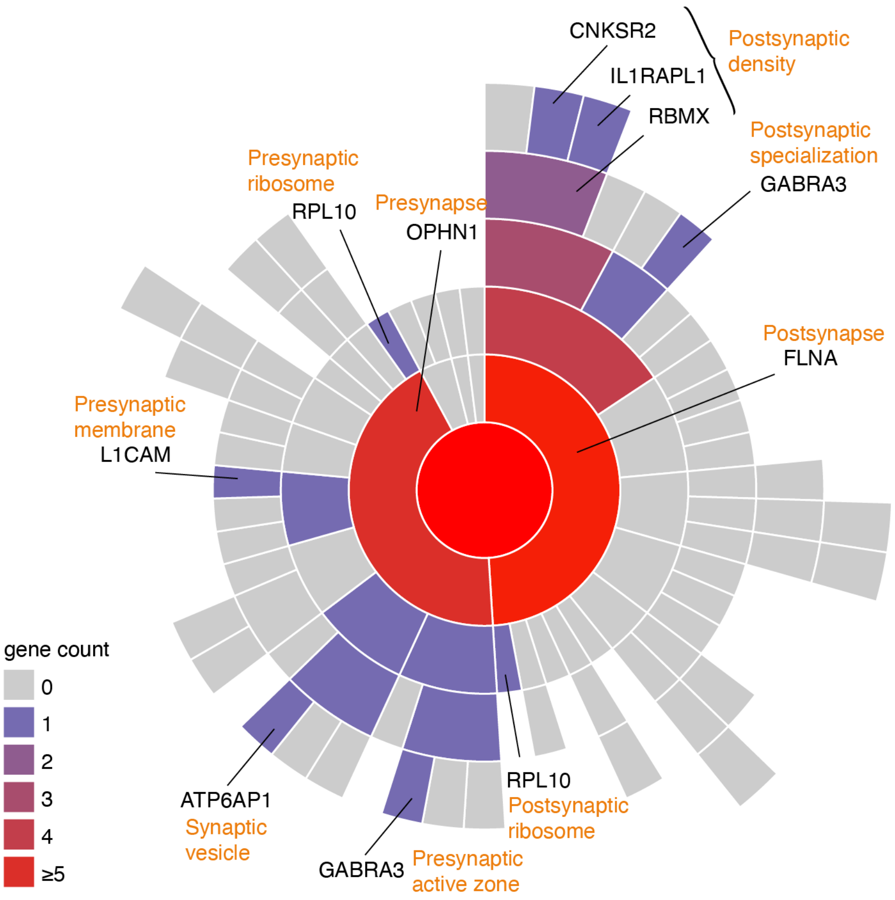


**Fig. S11: Mapping of all functionally mapped genes in FUMA to synaptic locations using SynGO.** Higher-level terms are in the center and child terms are in subsequent rings. The name of the term (in orange) and genes mapped to the term (in black) are annotated.

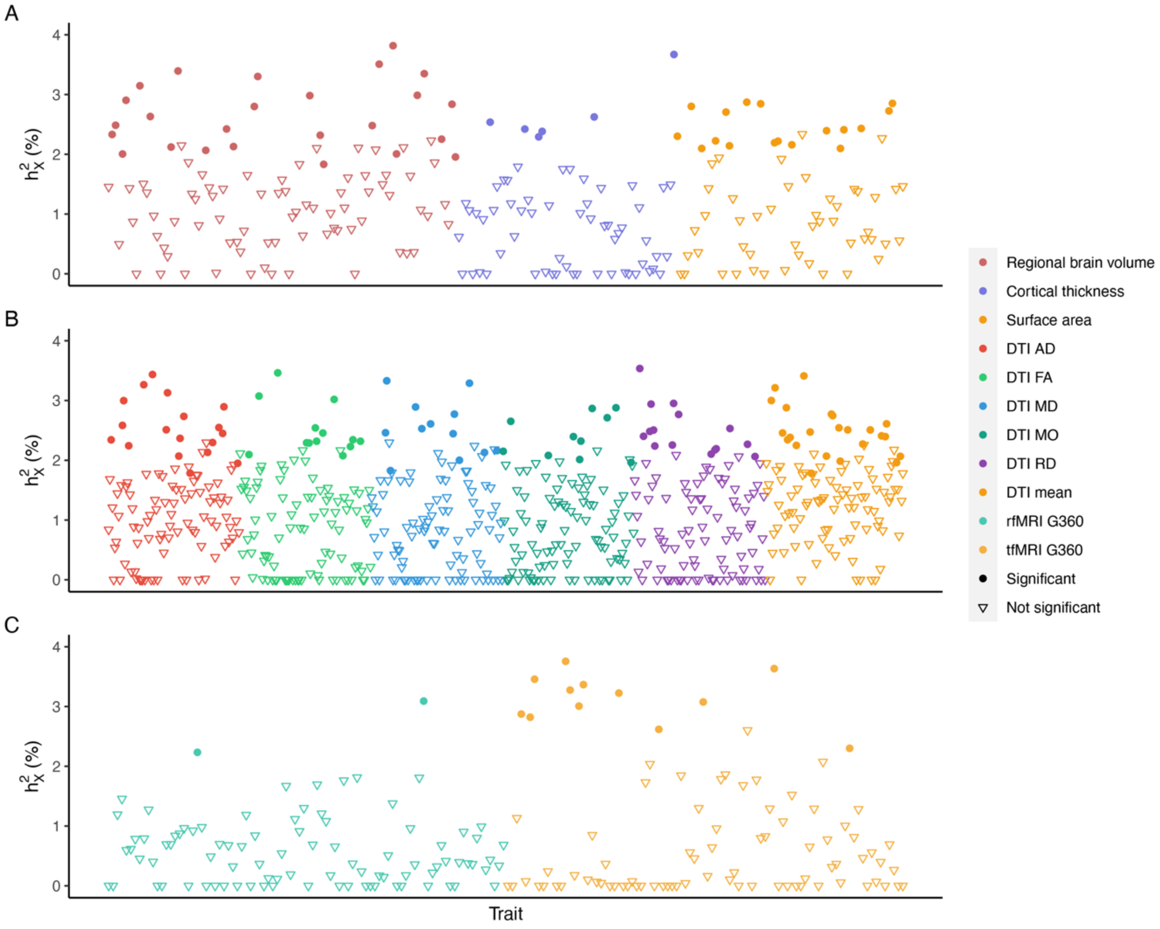


**Fig. S12: Estimated X-linked heritability ($h_X^2$) in males.** The different categories of traits are distinguished by color and traits with significant $h_X^2$ estimates are shown in solid circles. Insignificant traits are shown in inverted triangles. In total, 155 traits have significant signals. A) sMRI; B) DTI; and C) fMRI.

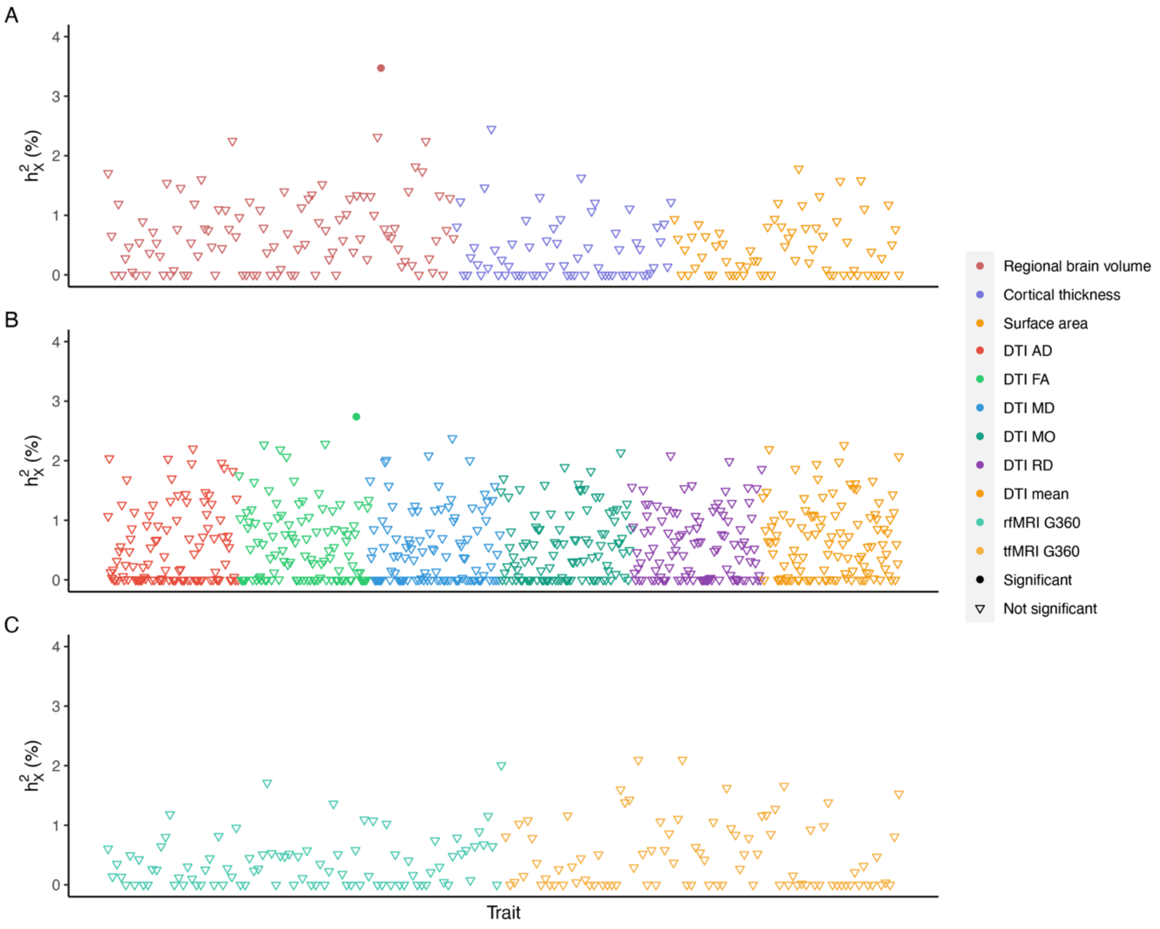


**Fig. S13: Estimated X-linked heritability ($h_X^2$) in females.** The different categories of traits are distinguished by color and traits with significant $h_X^2$ estimates are shown in solid circles. Insignificant traits are shown in inverted triangles. In total, only two traits have significant signals. A) sMRI; B) DTI; and C) fMRI.

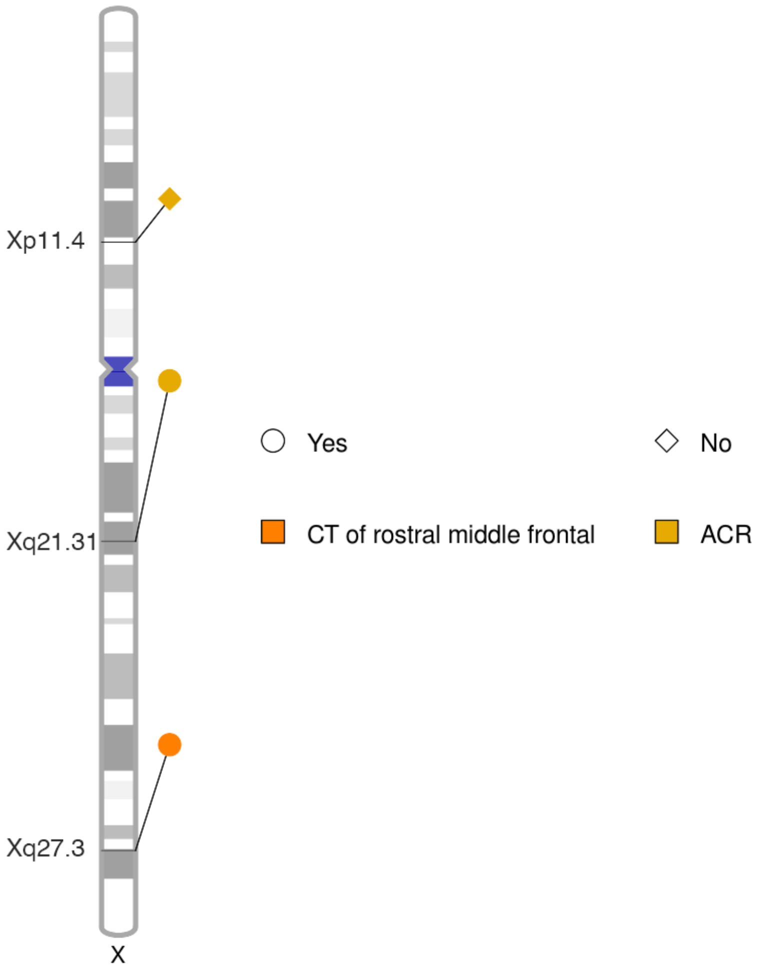


**Fig. S14: Genomic loci displaying significantly different effects between sexes.** Each trait is labeled with a different color. The name of each genomic region is annotated on the ideogram. “Yes” and “No” mean whether the signal can be recovered or not by a meta-analysis of sex-stratified results using Stouffer’s method (Methods).

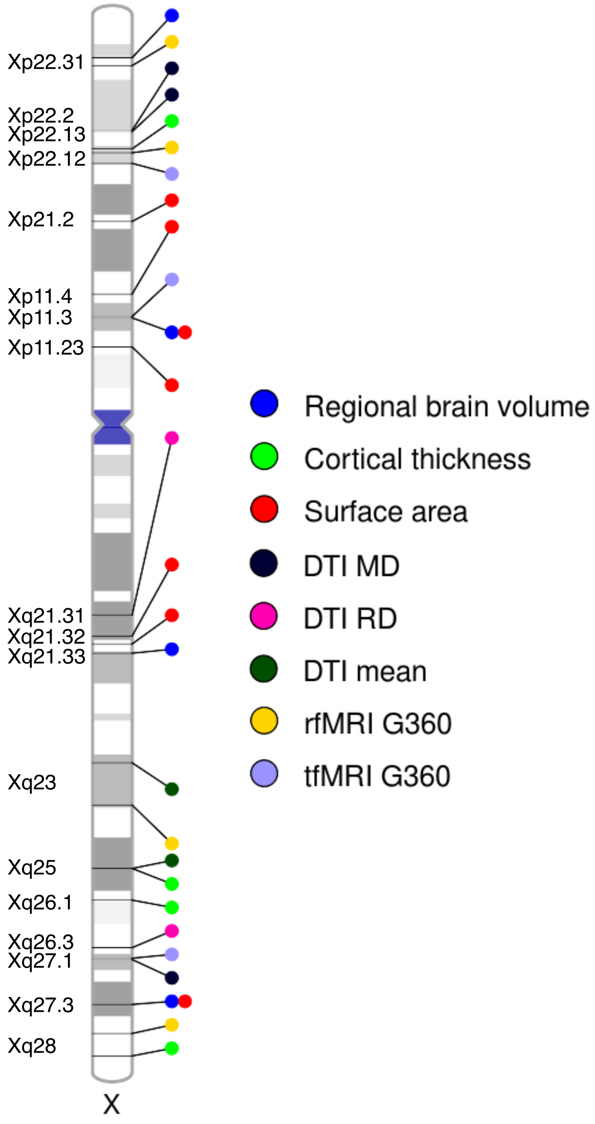


**Fig. S15: Genomic loci displaying significantly different profiles between males classified by phenotypic quantiles at distribution tails.** Each category of traits is labeled with a different color. The name of each genomic region is annotated on the ideogram.

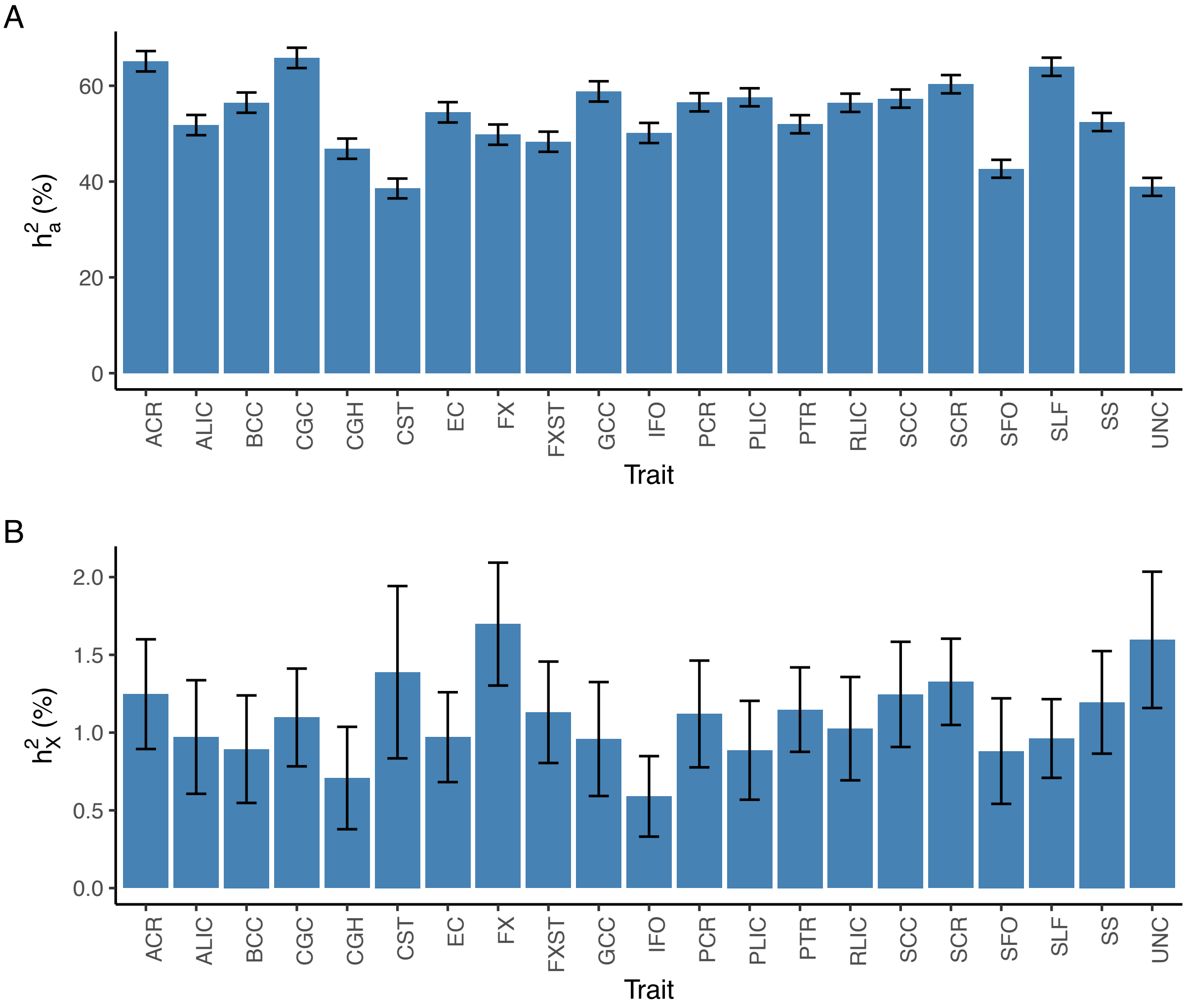


**Fig. S16: Heritability estimates of DTI tract-mean traits.** A) Autosomal heritability ($h_a^2$). B) X-linked heritability ($h_X^2$).

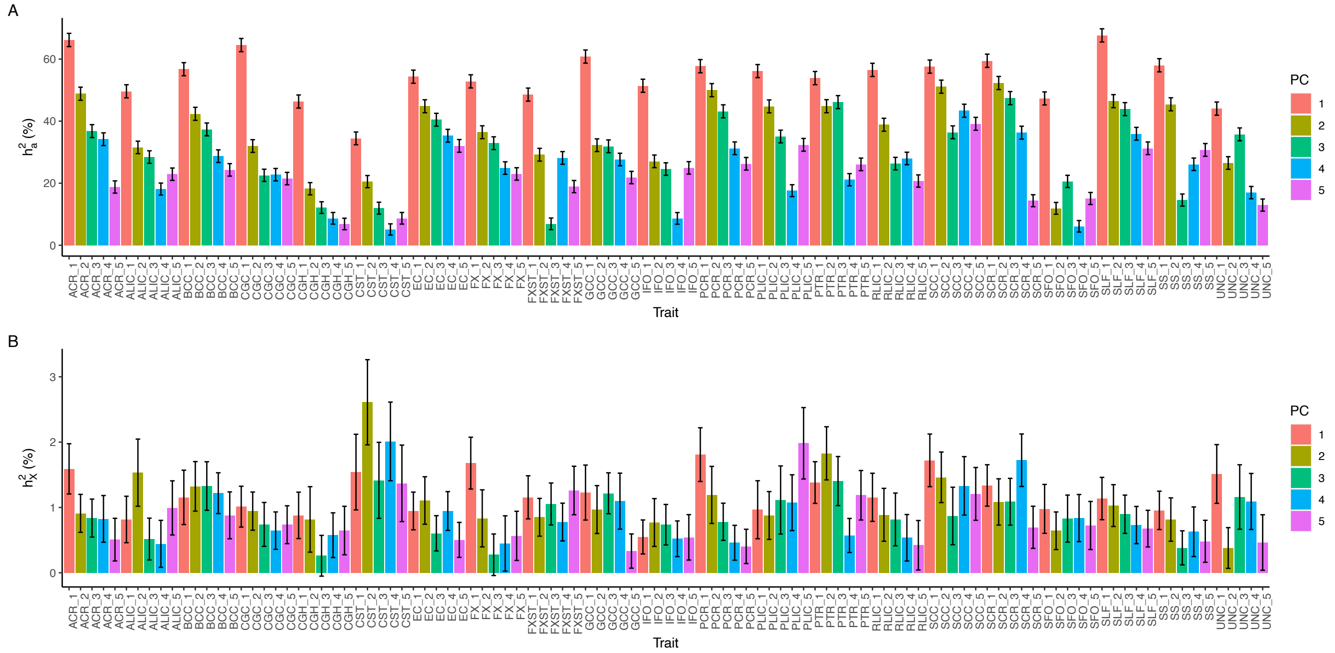


**Fig. S17: Heritability estimates of DTI PC traits.** A) Autosomal heritability ($h_a^2$). B) X-linked heritability ($h_X^2$).

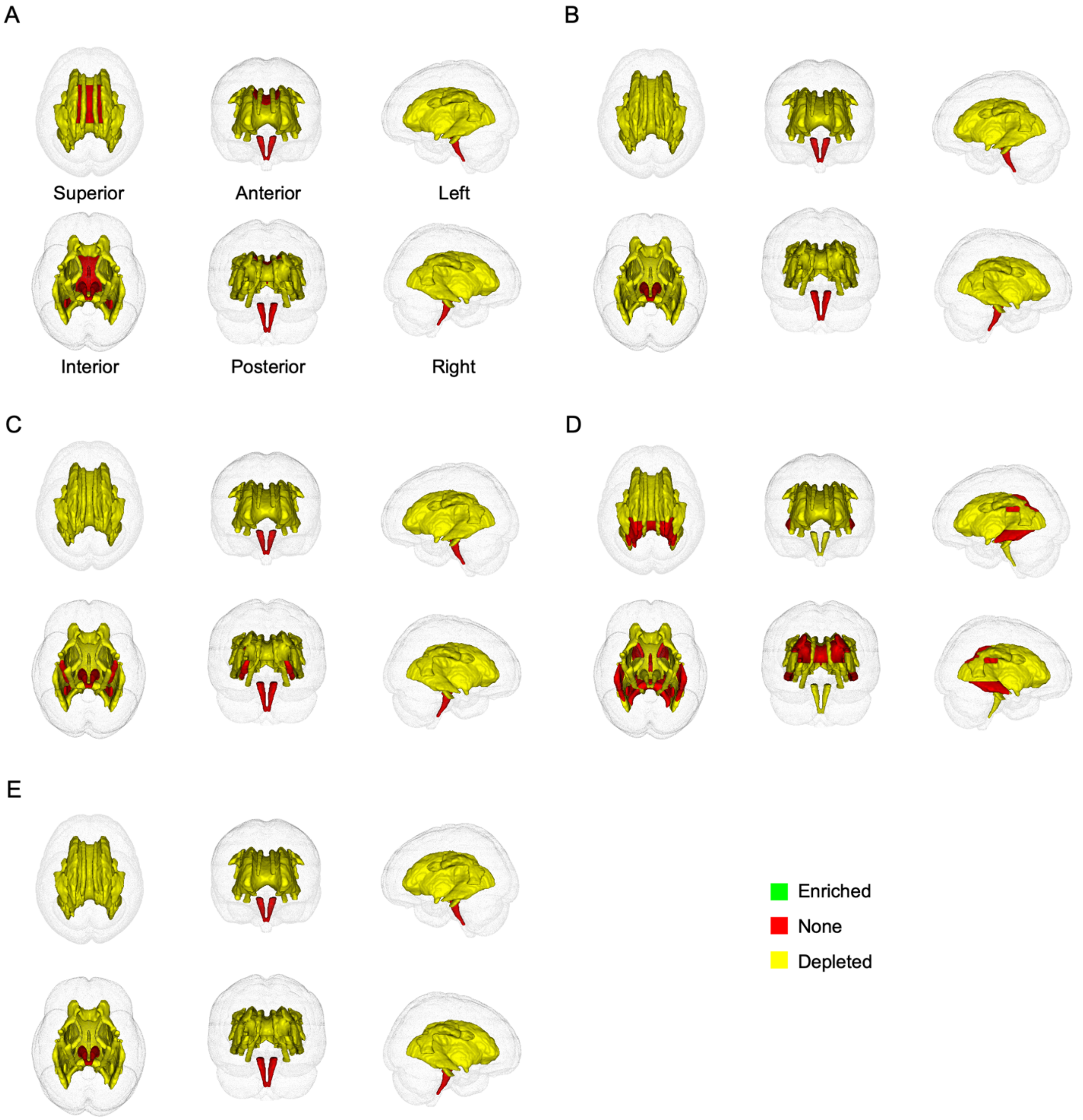


**Fig. S18: Atlases of enrichment of DTI tract-mean traits evaluated by different metrics.** A) AD; B) FA; C) MD; D) MO; and E) RD.

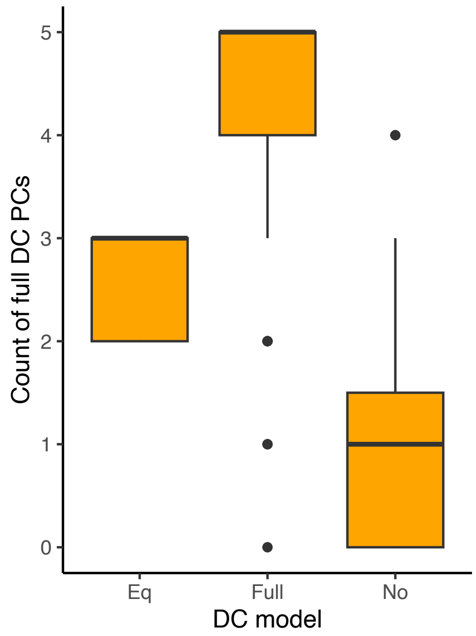


**Fig. S19: Consistency between the number of full DC PCs and the DC of tract-mean traits.**

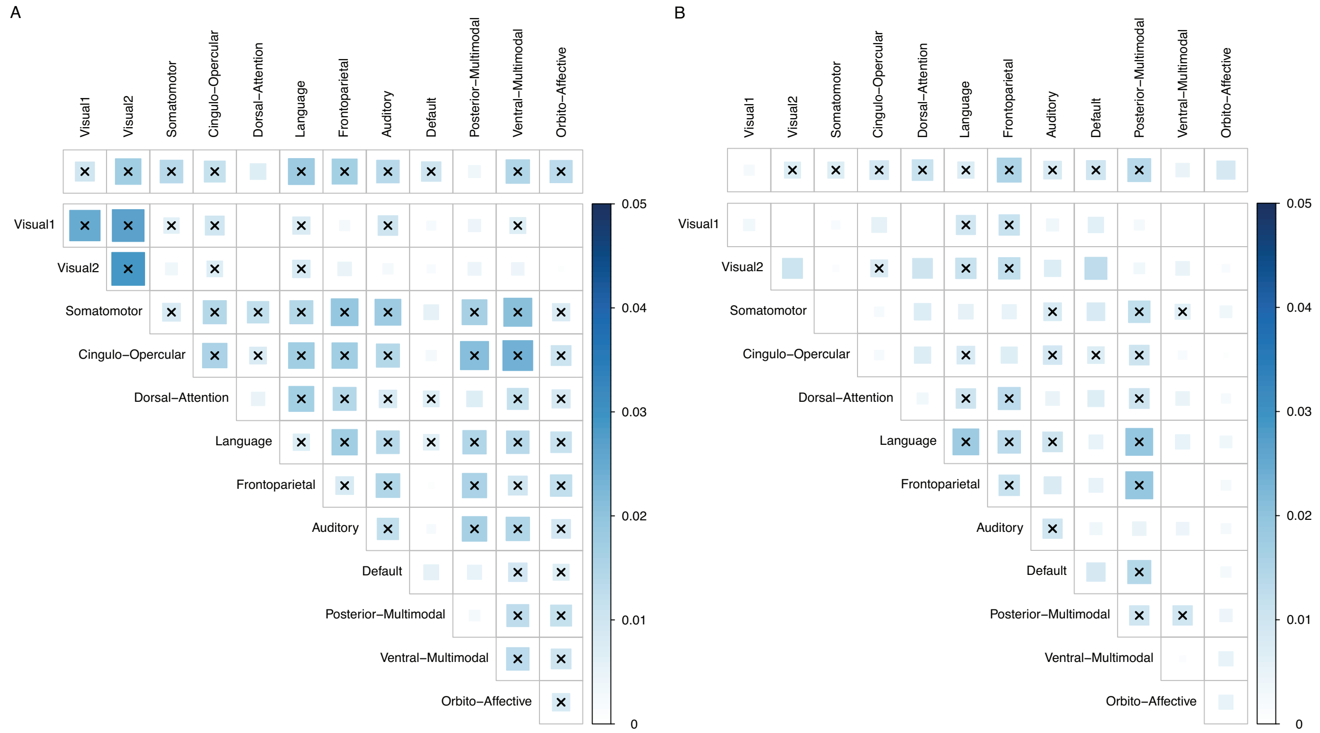


**Fig. S20: Heritability estimates ($h_X^2$) of fMRI traits.** The cells in upper triangle matrix are $h_X^2$ estimates of mean functional connectivity traits. The cells above the top margin are $h_X^2$ etimates of mean amplitude traits. Significant traits are annotated by asteroid. A) rfMRI. B) tfMRI.

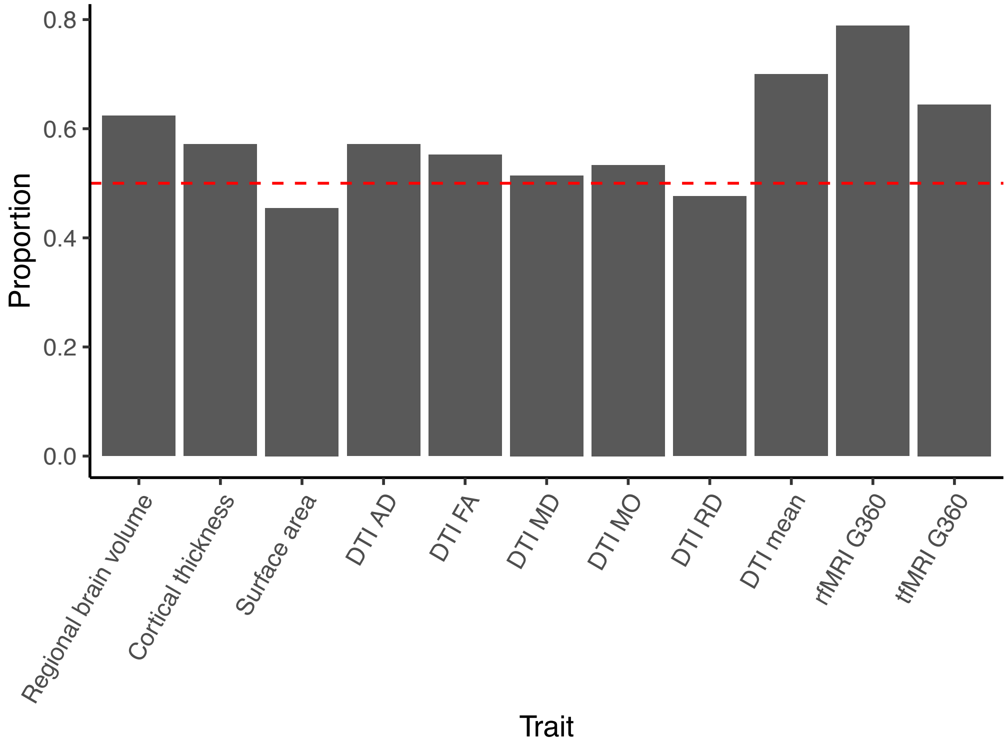


**Fig. S21: Proportion of traits where males have a larger phenotype than females.** The red dash line represents 0.5.

**Table S1.**

**Details for complex brain imaging traits.**
Sheet 1: This sheet shows trait categories and numbers of traits.
Sheet 2: This sheet shows abbreviations and full names for white matter tracts.
Sheet 3: This sheet shows DTI metrics and descriptions.
< in a separate .xlsx file >

**Table S2.**

**Results of dosage compensation and enrichment analysis for X-linked heritability.**
< in a separate .xlsx file >

**Table S3.**

**Basic information of XWAS.**
< in a separate .xlsx file >

**Table S4.**

**Independent significant SNPs identified by LD pruning.**
Sheet 1: Nominal significant SNPs (p-value < 5e-08) in LD > 0.6 were grouped under an independent significant SNP.
Sheet 2: We did a further round of pruning by restricting the adjusted p-values of identified independent significant SNPs (in Sheet 1) less than 5e-08, where the raw p-values were adjusted by wild bootstrap.
< in a separate .xlsx file >

**Table S5.**

**Significant loci identified by LD pruning.**
Sheet 1: Nominal significant SNPs (p-value < 5e-08) in LD > 0.6 were grouped under an independent significant SNP. The LD blocks defined by independent significant SNPs with distances less than 250kb with each other were merged into a single genomic locus.
Sheet 2: Based on results in Sheet 1, we further restricted that the top SNP for each locus was significant after adjusting for multiple hypothesis testing for all the traits using wild bootstrap. Whether the trait-locus pairs were newly identified and whether they were replicated by UKBE subjects only and by meta-analysis for UKBE, UKBA and UKBSAC subjects were also marked.
< in a separate .xlsx file >

**Table S6.**

**Significant variants of meta-analysis for sex-stratified XWAS results.**
Sheet 1: The meta-analysis was performed by METAL (https://genome.sph.umich.edu/wiki/METAL_Documentation). The raw p-values were adjusted by wild bootstrap considering the number of traits. The significant variants were those with adjusted p-values less than 5e-08.
Sheet 2: The bolded results are new SNPs that were not identified in the sex-agnostic XWAS.
< in a separate .xlsx file >

**Table S7.**

**XWAS replication results.** The top SNP for each locus was presented. A locus was replicated if the top SNP in the replication study was lower than the Bonferroni threshold (0.05/72 = 0.00069).
Sheet 1: The XWAS replication results using phase 4 European-ancestry subjects (UKBE, n = 4,181). Replicated associations are bolded.
Sheet 2: The XWAS replication results using phase 1-4 African-ancestry subjects (UKBA, n = 295).

Sheet 3: The XWAS replication results using phase 1-4 South Asian-ancestry and Chinese-ancestry subjects (UKBSAC, n = 462).
Sheet 4: The individual XWAS replication results were meta-analyzed using METAL (https://genome.sph.umich.edu/wiki/METAL). Replicated associations are bolded.
< in a separate .xlsx file >

**Table S8.**

**Significant variants of meta-analysis for XWAS discovery and white replication results.** The meta-analysis was performed by METAL (https://genome.sph.umich.edu/wiki/METAL_Documentation). The raw p-values were adjusted by wild bootstrap considering the number of traits. The significant variants were those with adjusted p-values less than 5e-08.
< in a separate .xlsx file >

**Table S9.**

**NHGRI-EBI GWAS catalog (2023.06) lookup for nominal significant SNPs (p-value < 5e-08) and significant SNPs after further adjustment.**
Sheet 1: The lookups in this sheet are based on nominal significant SNPs (p-value < 5e-08).
Sheet 2: The lookups in this sheet are based on significant SNPs after adjusting for multiple hypothesis testing for all the traits by wild bootstrap (adjusted p-value < 5e-08), as well as other SNPs that are in LD > 0.6 with these significant SNPs.
< in a separate .xlsx file >

**Table S10.**

**Independent significant SNPs mapped to genes based on expression quantitative trait locus (eQTL) mapping.** PAR SNPs had no results.
< in a separate .xlsx file >

**Table S11.**

**Summary data-based Mendelian randomization results.** The summary statistics of CAGE whole-blood eQTL analysis were provided by Sidorenko et al. (*44*). The analysis was conducted by SMR module in GCTA (https://yanglab.westlake.edu.cn/software/gcta/#Overview). We set all parameters as default.
< in a separate .xlsx file >

**Table S12.**

**Biological annotation for prioritized genes using DAVID Bioinformatics Database (https://david.ncifcrf.gov/home.jsp).**
< in a separate .xlsx file >

**Table S13.**

**Biological annotation for prioritized genes using SynGO (https://syngoportal.org/).**
< in a separate .xlsx file >

**Table S14.**

**Sex phenotypic differences in complex brain imaging traits.**
< in a separate .xlsx file >

**Table S15.**

**X-linked heritability estimated in sex-stratified analysis.** The heritability was computed by GCTA (https://yanglab.westlake.edu.cn/software/gcta/#Overview). The likelihood-ratio test (LRT) was conducted by comparing the full model including both the genetic relatedness matries (GRMs) of autosomes and of the X-chromosome, with the reduced model including the GRM of autosomes only. The raw p-values were adjusted for the number of traits by Benjamini-Hochberg procedure to control the false discovery rate (FDR) at 0.05 level.

< in a separate .xlsx file >

**Table S16.**

**Significant variants in sex-stratified XWAS.**

< in a separate .xlsx file >

**Table S17.**

**Significant variants in meta-analysis for sex-stratified XWAS using UKB discovery data and UKB replication data of white subjects.**

< in a separate .xlsx file >

**Table S18.**

**Variants that had significantly different effect sizes between sexes.** The significant variants were then meta-analyzed across sexes using the Stouffer's method.

< in a separate .xlsx file >

**Table S19.**

**Differences in genetic profiles between subjects classified by phenotypic quantiles.**

< in a separate .xlsx file >

**Table S20.**

**MAGMA gene-based analysis results.** Summary statistics of nominal significant variants (p-value < 5e-08) were utilized in the analysis. GRCh37 was used to map SNPs to genes by physical location.

< in a separate .xlsx file >

**Table S21.**

**Independent significant SNPs mapped to genes based on functional consequence.** The independent significant SNPs were in LD > 0.6 with the top SNP of each locus.

< in a separate .xlsx file >

**Table S22.**

**Mapped genes identified in functional mapping using nominal significant SNPs at 5e-08 level.**

< in a separate .xlsx file >

**Table S23.**

**Gene mapping using chromatin interaction profiles with H-MAGMA.**

< in a separate .xlsx file >
